# Repurposing of Glatiramer Acetate to Treat Heart Diseases

**DOI:** 10.1101/2023.09.30.23295943

**Authors:** Gal Aviel, Jacob Elkahal, Kfir Baruch Umansky, Hanna Bueno-Levy, Shoval Miyara, Daria Lendengolts, Lingling Zhang, Zachary Petrover, David Kain, Tali Shalit, Rina Aharoni, Ruth Arnon, David Mishaly, Uriel Katz, Dean Nachman, Mahdi Ammar, Rabea Asleh, Offer Amir, Eldad Tzahor, Rachel Sarig

**Author notes:** Corresponding authors’ (R.S.); (E.T.).

## Abstract

Myocardial injury may ultimately lead to adverse ventricular remodeling and development of heart failure (HF), which is a major cause of morbidity and mortality worldwide. Given the slow pace and substantial costs of developing new therapeutics, drug repurposing is an attractive alternative. Studies of many organs, including the heart, highlight the importance of the immune system in modulating injury and repair outcomes. Glatiramer-acetate (GA) is an immunomodulatory drug prescribed for patients with multiple sclerosis. Here we report that short-term GA treatment improves cardiac function and reduces scar area in a mouse model of acute myocardial infarction, as well as in a rat model of ischemic HF. We provide both *in vivo* and *in vitro* mechanistic evidence indicating that in addition to its immunomodulatory functions, GA exerts beneficial pleiotropic effects, including cardiomyocyte protection and enhanced angiogenesis, mediated partially by extracellular vesicles carrying a pro-reparative cargo. Finally, as GA is a widely used drug with established efficacy and safety history, we conducted a small, prospective, randomized trial to determine its effect on patients admitted to the hospital with acute decompensated HF (ADHF). Strikingly, a short-term add-on administration of GA, resulted in marked reduction in the cytokine surge and NT-proBNP levels, both associated with acute HF exacerbations. Overall, these findings demonstrate the efficacy of GA in attenuating acute myocardial injury and modulating the inflammatory process in animal models and humans and highlight the potential of GA as a future therapy for a myriad of heart diseases.

**One Sentence Summary:** Glatiramer acetate promotes reparative processes in rodent models of cardiac injury and reduces the inflammatory process in ADHF patients.

## INTRODUCTION

Heart failure (HF) is a leading cause of morbidity and mortality worldwide. While improvements in revascularization techniques and medical care have significantly reduced mortality rates from acute myocardial infarction (MI), the incidence of ischemic cardiomyopathy is increasing ^1^. The pathogenesis of HF is intricately related to a chronic inflammatory process, leading to irreversible loss of cardiomyocytes (CMs) ^2^. While a transient inflammation is essential for tissue healing, including cardiac repair ^3,4^, a maladaptive immune response may contribute to adverse left ventricle (LV) remodeling and development of chronic HF. The idea of using immunomodulatory drugs to ameliorate LV remodeling and improve heart function after acute MI has been tested in several clinical trials which yielded conflicting results^5^.

The development of new drugs is a laborious, long-term, and costly process ^6^. Hence, repurposing approved drugs to treat new indications is an appealing strategy for expanding the therapeutic armamentarium to treat patients. Glatiramer acetate (GA) is a synthetic random copolymer composed of four amino acids that has been used for years for the treatment of multiple sclerosis (MS) ^7^. Originally, GA was designed to resemble the autoantigen myelin basic protein and shown to have beneficial effects in animal model of MS, experimental autoimmune encephalomyelitis ^8^, yet multiple studies revealed its broad immunomodulatory and anti-inflammatory mechanism of action, at different levels of both the innate and the adaptive immune responses ^9,10^. As such, GA binds promiscuously to major histocompatibility complex (MHC) molecules, acting both as an MHC blocker ^11^ and a T cell receptor antagonist ^12^, leading to inhibition of pathological effector functions. GA has also been shown to modulate the properties of dendritic cells and monocytes to preferentially stimulate type 2 helper T (Th2) cell-like responses ^13^, inducing the secretion of anti-inflammatory cytokines ^14,15^ and elevation of T-regulatory cells (Tregs) ^16^. GA-induced Th2 cells and Tregs were shown to accumulate in the injury site and secrete *in situ* anti-inflammatory cytokines and growth factors that suppress the inflammation and augment repair processes ^17,18^. Based on its broad immunomodulatory mode of action, potential applications of GA for additional pathologies were investigated, showing its beneficial effects in prevention of immune rejection ^19^, improvement of stem cells engraftment ^20^, amelioration of inflammatory bowel disease (IBD) ^21,22^, and repair of liver fibrosis ^23^. Due to the immunomodulatory mechanism of action of GA, its efficacy in various pathological systems, and its broad safety profile, we hypothesized that it might mitigate the pathological inflammatory process associated with myocardial injury, thereby preventing its progression, and improving cardiac outcomes.

In the present work, we initiated a drug repurposing process of GA for the treatment of heart diseases from the preliminary pre-clinical work in murine models of acute myocardial ischemia to the completion of a phase 2a clinical trial in patients with HF. First, we tested the potential application of GA for improving heart function using murine models of myocardial ischemia. Our data show beneficial pleiotropic effects of GA treatment on the injured heart, manifested by improved cardiac function and reduced scar area in a mouse model of acute MI as well as in a rat model of HF. Examination of GA’s mode-of-action revealed that in addition to its known immunomodulatory effects, shown here for the first time in the context of myocardial inflammation, it promoted CM protection from ischemia-induced death, reduced fibrosis and enhanced angiogenesis. Administration of extracellular vesicles (EVs) isolated from GA-treated hearts recapitulated GA’s effects in a mouse model of acute MI, suggesting that these beneficial paracrine effects were mediated by EVs. *In vitro* and *ex vivo* assays further supported a paracrine effect of stromal cells on CMs.

Next, we proceeded to conduct a small phase 2a, open-label, randomized-controlled clinical trial to assess the potential and safety of GA treatment in patients hospitalized with ADHF. GA therapy added to standard HF therapy resulted in a significant blunting of the cytokine surge associated with ADHF and improvement in HF severity, as represented by a remarkable reduction in the levels of the natriuretic peptide NT-proBNP, compared to standard HF therapy alone. Overall, our data reveal novel protective and reparative effects of GA in rodent models of cardiac ischemia, as well as its safety and beneficial effects in ADHF patients. Collectively, these results highlight the potential of repurposing GA as a future therapy for patients with heart diseases.

## RESULTS

### Transient treatment with GA results in improved cardiac function after acute MI

Searching for drug candidates that could be repurposed to treat heart diseases, we postulated that the beneficial immunomodulatory effects of GA could be harnessed to promote myocardial repair. To test this hypothesis, we subjected adult mice to acute MI^24^ and divided into two groups, receiving daily intraperitoneal (i.p) injections of either GA or solvent as a control for 14 days, starting from the day of surgery (Fig. 1A). While transthoracic echocardiography confirmed the presence of a significant injury in both groups 2 days post-injury (dpi), at 35 dpi, a significant improvement in systolic function was apparent only in the GA-treated group, as displayed by both ejection fraction (EF) and fractional shortening (FS) (Fig. 1B,C). Quantification of the scar area in histological sections showed a significant reduction in infarct size following GA treatment (Fig. 1D,E). In addition, stratification of scar area according to size revealed that large scars, encompassing more than 30% of LV volume, were present only in the control group. GA treatment yielded similar results also when calculating ischemic area-at-risk and infarct zone at 4 dpi (Fig. 1F,G), excluding the possibility of surgical inconsistencies in the findings and ensuring the quality control of the procedure.

**Figure 1:**
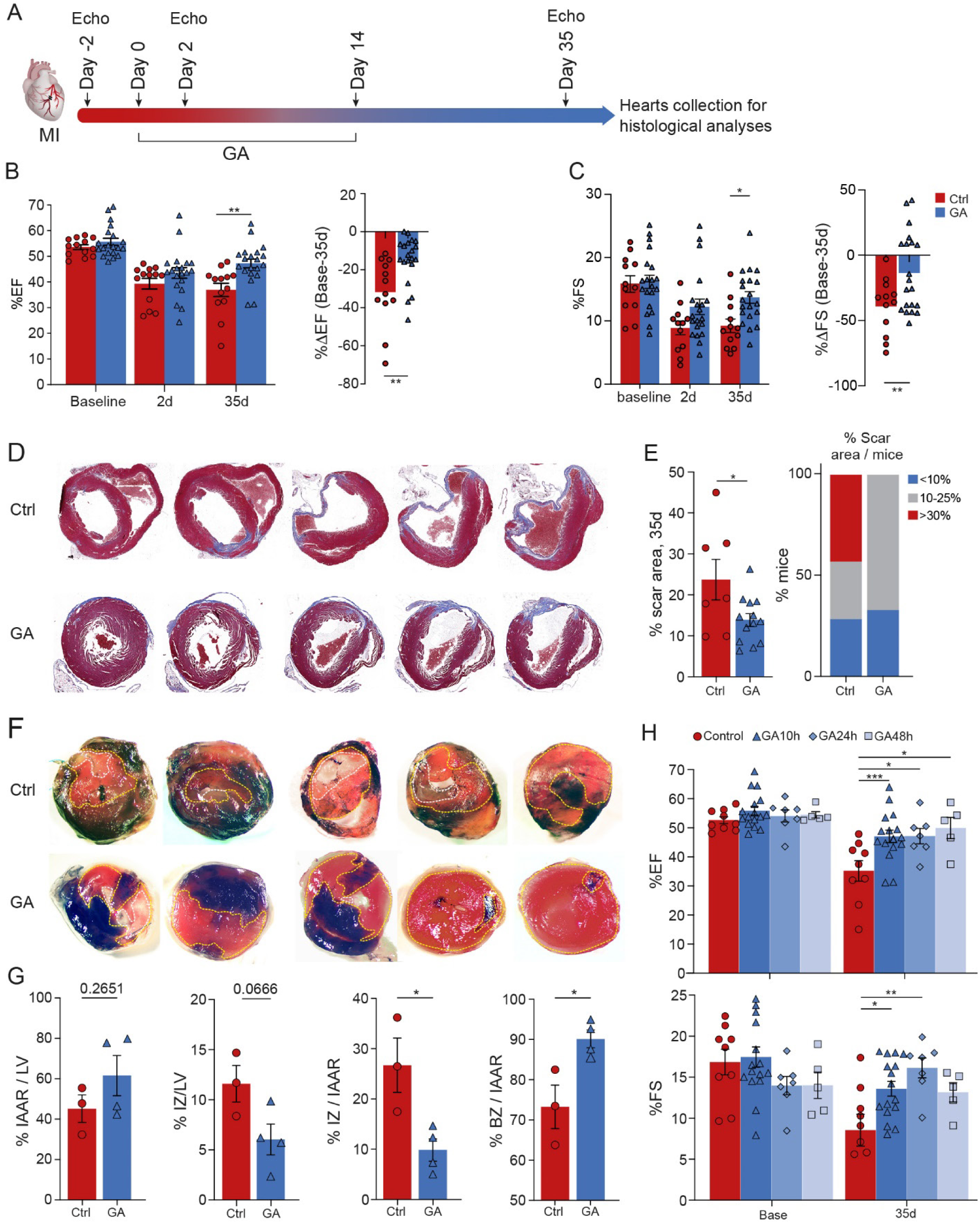
Glatiramer acetate improves mouse cardiac function after myocardial infarction. **A,** A scheme of the experimental procedure. Mice were divided into two groups receiving either GA (2 mg/animal/day) or control (PBS / mannitol) by daily IP injections, from day 0 (day of MI) up to 14 days post-MI. Echo measurements were performed at baseline (2 days before MI) and at 2-, 14- and 35-days post-MI. Histological analysis was performed postmortem. **B,C,** Echocardiographic measurements of ejection fraction (EF, **B**) and fractional shortening (FS, **c**) at baseline, 2 days and 35 days after panels). The percent reduction in EF or FS is shown on the right (n_control_ = 13, n_GA_ = 20). **D,** Masson’s trichrome staining of heart sections derived from representative PBS-(Ctrl, top) or GA-treated hearts (bottom). **E,** Left: Mean percentage of scar area in sections from GA- or control-treated hearts 35 days after MI. Right: Scar stratification according to size reveals that large scars (>30% of LV) were found only in the control group. **F,** representative sections of control (upper row) and GA-treated (lower row) hearts, showing well perfused myocardium (black), IAAR (dashed *yellow* line) and IZ (dashed *white* line). **G,** Quantification of % IAAR/LV showing no significant differences between the groups, while % IZ/IAAR and %BZ/IAAR demonstrate a significant reduction in IZ following GA treatment and increase in BZ, suggesting myocardial protection (n_GA_ = 4, n_control_ = 3). IAAR = ischemic area-at-risk, IZ = infarct zone, BZ = border zone. **H,** A wide temporal therapeutic window for GA is shown by echo measurements of EF (upper) and FS (lower) in animals treated with PBS (control) or GA at the day of injury, 24- or 48-hours post-MI. n_control_ = 9, n_GA/t0_ = 16, n_GA/24h_ = 7, n_GA/48h_ = 5.

Subsequently, we tested whether the therapeutic window for GA administration might be extended, which is relevant for the treatment of late-arrival MI patients^25^. To that end, mice were subjected to MI and treatment was postponed for 24- or 48-hours post-MI. Results showed that delayed administration of GA improved systolic function to a similar extent as compared to immediate treatment (Fig. 1H). This suggests a wide temporal window for GA administration, relevant for late arrival patients.

### GA treatment attenuates acute inflammatory response and promotes a pro-reparative immune phenotype after acute MI

Following acute MI, the ischemic myocardium activates an intense inflammatory response, mostly of the innate immune system, followed by infiltration of neutrophils and bone marrow-derived monocytes. The early dominance of innate immunity is then replaced by an adaptive immune response ^26^. Based on the known immunomodulatory effects of GA in various models of organ injury ^23,27,28^, we postulated that it would inhibit the acute inflammatory response following MI and promote a pro-reparative immune phenotype. To gain deeper insights on the effects of GA on the inflammatory response following acute MI, we performed single-cell expression profiling on CD45+-enriched cardiac cells at 1 and 4 dpi, using the 10x Genomics Chromium platform. Adult mice were subjected to acute MI and treated daily with either GA or control until their sacrifice (Fig. 2A). Echocardiography performed at 3 dpi demonstrated a significant beneficial effect in the GA-treated group on %EF already at this early timepoint, indicating a potential protective effect of GA on the ischemic myocardium (Fig. 2B). Transcriptional profiles of 49,369 cells were captured after quality control filtering. Distinct cell populations, represented by a total of 19 clusters, were visualized in uniform manifold approximation and projection (UMAP) reduced dimensionality plots (Fig. 2C and Fig. S1A). Cluster annotation was performed using canonical markers for each cell type (Fig. 2C and Methods).

**Figure 2:**
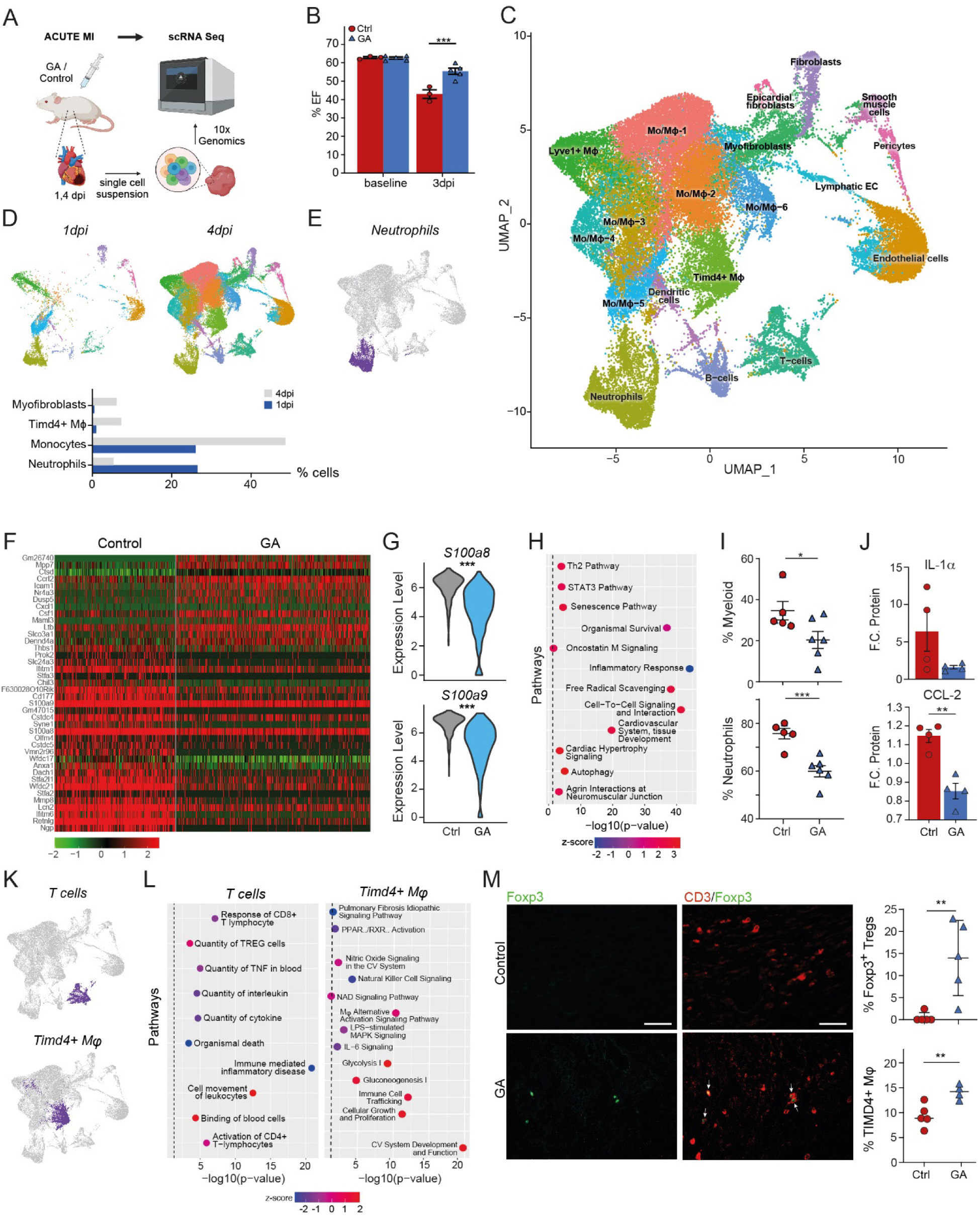
GA treatment promotes a pro-reparative immune phenotype. **A,** Experimental design. Sample size – 1 dpi (n_GA_ = 1, n_Control_ = 1), 4 dpi (n_GA_ = 2, n_Control_ = 2). **B,** EF values at baseline and 3 dpi, demonstrating a cardiac protective effect induced by GA treatment. **C,** Dimensional reduction with UMAP. **D**, Frequency plots of specific cell types according to days. **E**, Neutrophilic cluster, **F,** Heatmap of DEG between GA and control at 1 dpi. Adjusted *P*-value <0.05. **G**, A violin plot for canonical markers for neutrophil activation. **H**, IPA performed on DEG of neutrophil cluster at 1 dpi. **I,** FACS analysis measuring the percentage of myeloid cells and neutrophils in control versus GA-treated mice 24h after MI (n_control_ = 5, n_GA_ = 6). **J**, Levels of pro-inflammatory mediators IL-α and CCL-2 were measured 24h post-MI in serum by ELISA. Levels are shown as fold-change from uninjured mice. n_GA_ = 4, n_mannitol_ = 4, n_sham_ *=* 3. **K**, highlights of T cells and Timd4+ resident macrophages. **L**, IPA performed on DEG of T cells and Timd4+ macrophages on respective clusters of CD45+ cell subset (adjusted *P* value <0.05). **M**, *upper panel*: FACS analysis of Timd4+ macrophages in GA-treated hearts 4 dpi (n_mannitol_ = 5, n_GA_ = 4) and Foxp3+ Tregs in GA-treated hearts 7 dpi (n = 5 for each group). *Lower panel*: Representative heart sections derived from GA- or PBS-treated mice 7 dpi that were co-immunostained with anti-CD3 and anti-Foxp3. Scale bars: 60µm.

Neutrophils are the first immune cells to infiltrate the myocardium in response to injury, peak at day 1 and quickly undergo apoptosis thereafter ^29^. As expected, neutrophils were the predominant immune cells detected at 1 dpi (∼26.3% of total cells), and by day 4 their numbers substantially decreased to 5.6% of total cells (Fig. 2D), validating the injury. The monocytic/macrophage population demonstrated opposite kinetics, comprising 26% of total cells in 1 dpi, and increasing to 48.5% by day 4. Of note, at 1 dpi the majority of *Cd68+* monocytes were *Lyve1+Timd4+* resident cardiac macrophages, which were further increased at 4 dpi due to the accumulation of *Lyve1-Timd4+* macrophages (Fig. 2D). Another validation for the injury can be shown by the activation of *Col1a1+Postn+* myofibroblasts that increased by 12-fold at 4 dpi (Fig. 2D).

The average abundance of T- and B-cells was largely constant (∼4% each, not shown), consistent with the known slower kinetic of adaptive immunity ^30^. Excessive neutrophil activation has been associated with increased infarct sizes due to the increased tissue damage ^31^ and with occurrence of fatal ventricular fibrillation ^32^. We identified a distinct neutrophil cluster based on canonical markers (Fig. 2E). Differential gene analysis comparing the GA and Control groups at 1 dpi demonstrated 726 differentially expressed genes (DEG) as well as down-regulation in neutrophil activation markers such as *S100a8* and *S100a9* in the GA group (Fig. 2F-G). These genes bind to Toll-like receptors and activate inflammasome-dependent pathways, thereby intensifying inflammatory activation, resulting in increased damage ^33^. Ingenuity pathway analysis (IPA) of DEG demonstrated in the GA treated mice down regulation in pathways related to *Inflammatory response*, and upregulation in pathways related to *Free radical scavenging*, *Senescence*, *STAT3* and *Autophagy,* associated with neutrophil silencing and clearance from the tissue ^34^ (Fig. 2H). To substantiate these findings, we analyzed neutrophil myocardial infiltration at 1 dpi using FACS, which demonstrated that GA administration had resulted in a significant reduction in their numbers (Fig. 2I and Fig. S1B). The attenuation of the inflammatory response at 1 dpi was further demonstrated by the significant reduction observed in serum levels of pro-inflammatory cytokines induced by GA treatment (Fig. 2J).

Th2 cells and Tregs play an essential role in the resolution phase of tissue injury and in promoting repair processes in various tissues, including the heart ^35^. The immunomodulation activity of GA in patients with MS includes the stimulation of Th2-like responses ^13^, with concomitant elevation of Tregs ^16,36^. We detected a distinct cluster of *Cd3e+Cd3d+* T cells (Fig. 2K). IPA performed on the DEG between GA and control at 4 dpi revealed down-regulation in numerous pathways related to immune activation and upregulation in a pathway related to Treg cells (Fig. 2L). Immunostaining and flow cytometry demonstrated elevation in Tregs levels in GA-treated hearts (Fig. 2M and Fig. S1C), indicating that in addition to the immediate effect of GA on the pro-inflammatory response, it can also modulate the immune response in the injured heart towards reparative inflammation.

In the regenerative murine neonatal heart, tissue-resident macrophages expand and dominate the injured area, resulting in reduced inflammation, enhanced angiogenesis, and CM proliferation ^8^. However, in adult mice, this resident macrophage population is replaced, or outnumbered, by monocyte-derived macrophages that are prominently pro-inflammatory ^8^. Thus, coordinated temporal activation of distinct macrophage populations might be essential for cardiac healing. We detected a cluster of *Timd4+* resident cardiac macrophages that expanded in size by 4 dpi (Fig. 2K lower panel). IPA performed on the DEG between GA and control at 4 dpi revealed metabolic signatures favoring *Glycolysis*, *Gluconeogenesis* and *cellular growth and proliferation*. In addition, we noted a down regulation in pathways related to *Il-6 Signaling* and *natural killer cell signaling*, which could reflect a reparative phenotype induced by the Timd4+ resident macrophages (Fig. 2L). To corroborate this, we followed the dynamics of cardiac macrophage profiles at 4 dpi using FACS and detected a significant increase in the levels of TIMD4+ resident cardiac macrophages that limit adverse remodeling after MI ^37^ in the hearts of GA-treated mice (Fig. 2M).

Taken together, our data support an early anti-inflammatory effect induced by GA treatment. This is demonstrated by transcriptomic changes indicating reduced neutrophil activation, reduced myocardial infiltration of neutrophils, as well as by reduction in serum pro-inflammatory cytokines. Additionally, we provide evidence that GA induces a pro-reparative immune phenotype, reflected by transcriptomic changes seen at single cell level of T cells and cardiac resident macrophages, which are further supported by FACS and immunofluorescence analyses.

### Single cell profiling suggests multiple effects on endothelial and fibroblasts induced by GA treatment following MI

The single cell suspensions for the scRNA-seq experiment were enriched for CD45+ to mainly focus on the myocardial immune response following GA treatment. As expected, the samples also contained 11708 non-immune cells (mainly endothelial cells (EC) and cardiac fibroblasts (CF)). To gain better insight on these populations, we re-clustered them and generated an unbiased targeted UMAP for each population (Fig. S2A and Fig. 3A-C). The ECs consisted of 6 different clusters: lymphatic, venous, arterial, and 3 distinct capillary endothelial clusters (Fig. S2A). IPA performed on the DEG at 1 dpi of the general endothelial cluster demonstrated enrichment in pathways related to *Vasculogenesis*, *cell survival* and *cell movement* and down regulation in pathways related to *cell death of endothelial cells* in the GA group compared to control. This reflects a possible protective and proangiogenic effect induced as early as 1 day following GA treatment (Fig. S2B). Capillary sub-clustering consisted of Capillary-1 and Capillary-2 were detected at 1 dpi whereas Capillary-3 emerged only at 4 dpi (Fig. S2C). A Frequency plot revealed that the GA-treated group was enriched for Capillary-1, while Capillary-2 was more prevalent in the control group (Fig. S2D). Further pathway analysis demonstrated that Capillary-1 cluster is enriched with pathways related to *blood vessel morphogenesis* and various *angiogenesis* pathways, whereas the Capillary-2 cluster was enriched with inflammatory pathways, including pathways related to *interleukin-1 beta production* and *leukocyte cell-cell adhesion*, further supporting an angiogenic effect of GA (Fig. 2SE-F).

**Figure 3:**
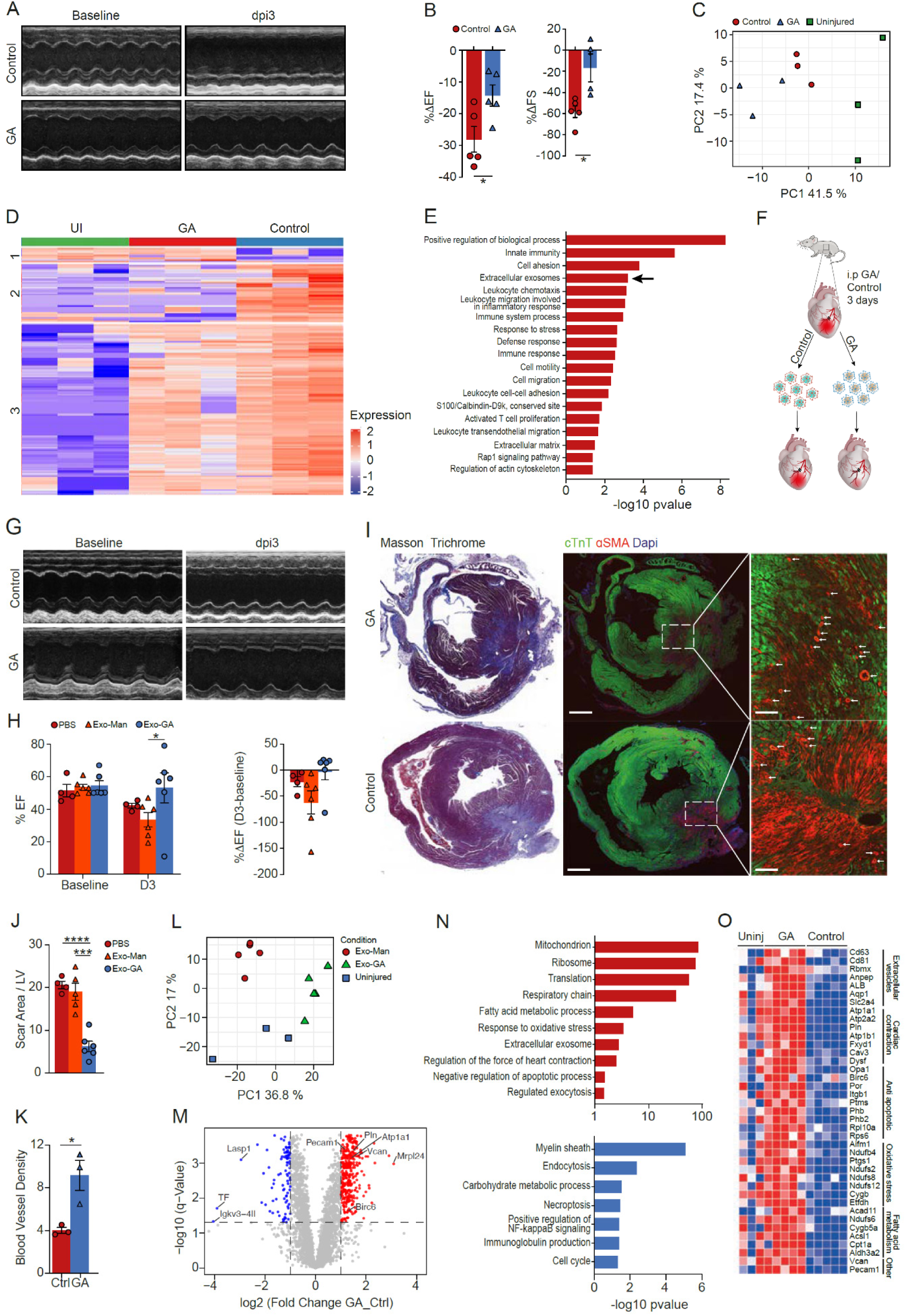
The protective effect of glatiramer acetate is mediated by extracellular vesicles. **A,** Representative M-mode echocardiography images, at 3 dpi. **B,** Analysis of echocardiographic systolic parameters. **C,** PCA of proteomic data. **D,** Heatmap of differentially expressed proteins (n=3 in each group). **E,** Gene ontology terms and pathway analysis. Arrow points to extracellular exosomes pathway. **F,** A graphical illustration of EV experiments (n=16). **G,** Representative M-mode echocardiography images, at 3 dpi. **H**, EF measurements (left), comparing the three experimental groups at baseline and 3 dpi. The right panel shows the change in EF as a fold-change from baseline levels. **I**, Masson Trichrome representative cardiac sections (left) and IF images (right) demonstrate smaller infarcts with large areas of viable myocardium, as well as higher density of αSMA+ blood vessels (arrowheads) in mice treated with GA-derived EVs. Scale bars: left panels, 750μm; right panels, 120μm. **J,K,** Analysis of infarct size (n=15) (**J**), and blood vessel density (n=6) (**K**) at 4 dpi. **L**, Proteomic analysis of cardiac EVs. Unsupervised PCA showing 3 distinct clusters of the experimental groups. **M**, ANOVA results of differentially expressed proteins between GA-derived and control-derived EVs. **N**, Gene ontology terms and pathway analysis, ranked according to −log10(corrected-p-value). **O**, targeted heatmap of individual proteins comprising key differentially changed pathways. Statistical significance was calculated using a two-tailed *t*-test, or ANOVA with Tukey’s correction for multiple comparisons, as appropriate.

The CF UMAP consisted of a distinct cluster of myofibroblasts and quiescent fibroblasts, 2 clusters of pericytes, and a distinct monocytic cluster (Fig. S3A). The monocytic cluster that was found in the targeted UMAP of both fibroblasts and endothelial cells expressed both monocytic gene markers as well as endothelial- or fibroblasts-markers, probably reflecting stromal cell phagocytosis by recruited monocytes/macrophages (Figs. S2A and S3B). As expected, these monocytes were absent at 1 dpi and appeared only at 4 dpi, reflecting the known kinetics of monocytic recruitment to the tissue. Pathway analysis of DEG revealed an intense activation of fibroblasts at 1 dpi in the GA-treated group compared to control, with enrichment of pathways related to *Wound Healing Signaling* (Fig. S3D, left). The analysis at 4 dpi demonstrated an opposite expression pattern, characterized by profound silencing of fibroblast activation, as evidenced by down regulation in pathways related to *Wound Healing*, and various pathways of *Fibrosis* (Fig. S3D, right). These data suggest that GA might induce a biphasic fibroblast activation response, characterized by an early acute activation, required for the immediate response, followed by a profound inactivation, that could reduce the deposition of collagen and the progression of fibrosis.

Overall, these data suggest that GA affects the transcriptome of cardiac ECs and fibroblasts. In fibroblasts it induces a biphasic activation and silencing pattern, and in ECs it promotes the protection and angiogenic responses.

### Extracellular vesicles isolated from GA-treated hearts carry a pro-reparative cargo and can recapitulate the beneficial effects of GA after acute MI

Next, we explored the impact of GA therapy on the proteomic landscape at 4 dpi. Here too, echocardiography of injured mice performed at 3 dpi indicated that the decline in systolic parameters was mitigated in GA-treated mice, indicating a potential protective effect of GA on the ischemic myocardium (Fig. 3A,B). Principle component analysis (PCA) performed at 4 dpi revealed three distinct clusters according to the treatment (uninjured, MI-control, MI-GA) (Fig. 3C). A total of 93 DE proteins were detected in the control group and 178 in the GA group (Fig. 3D). Pathway analysis of the DE proteins showed enrichment in pathways of innate immunity, leukocyte chemotaxis, T cell proliferation, and cell motility and migration in the GA treated hearts, all consistent with substantial modulation of the immune response (Fig. 3E). In addition, we noted a significant enrichment in proteins related to extracellular vesicles (EVs) (Fig. 3E). EVs are membrane-containing organelles, of 30-120 nm in diameter, which are secreted through exocytosis, carrying distinct cargo capable of exerting a myriad of effects, including repair ^38,39^. Additionally, the protective effect of endothelial cell-derived EVs was recently shown in a human heart-on-chip model ^40^, suggesting that GA could act by paracrine signaling mediated by EVs.

We therefore examined whether cardiac EVs derived from GA-treated animals can recapitulate the beneficial effects of GA on cardiac function after injury. For that, we isolated cardiac EVs from mice subjected to acute MI (Fig. S4A-D). Based on the proteomic analysis results, animals were treated for 3 days with either GA or control and on day 4 EVs were isolated from the LV and were administered immediately after LAD ligation to a second cohort of mice (Fig. 3F). Consistent with the proteomic data, a significant reduction in %EF values was apparent in animals that received a single intramyocardial injection of control-derived EVs or PBS (Fig. 3G,H). Notably, the reduction in systolic function was blunted by administration of EVs derived from GA-treated mice (Fig. 3G,H). In addition to functional cardiac improvement, histological analysis revealed a significant reduction in infarct size following GA-derived EV treatment, compared to control-treated hearts (Fig. 3I,J). As myocardial perfusion is a major determinant of cardiomyocyte death following acute ischemia, we assessed arterial density in the infarct zone. A significant increase in αSMA+ blood vessels and in CD31+ cells was observed in mice that were treated with GA-derived EVs compared to control EVs at 4dpi (Fig 3I-K, Fig S5), suggesting that GA-derived EVs exert a protective effect on myocardial coronary arteries. The strong αSMA+ staining observed inside the scar region, mainly in control hearts, reflects activated myofibroblasts. Taken together, our results suggest a protective effect of GA-derived EVs on CMs and coronary vasculature.

To analyze the differential protein content of cardiac EVs derived from GA- or control-treated hearts, we performed proteomic analysis of these vesicles. PCA revealed three distinct clusters: GA-derived, control-derived EVs, or EVs from uninjured hearts (Fig. 3L), indicating GA-dependent differences. Overall, 455 proteins were found to be significantly changed in the GA-derived EVs (Fig. 3M). GA-derived EVs exhibited significant enrichment in numerous gene ontology (GO) terms, such as negative regulation of apoptotic processes, cardiac contractility, respiratory chain, mitochondrion, and ribosome (Fig. 3N). A targeted heatmap based on the individual proteins that comprise these pathways further highlighted distinct proteins in GA-derived EVs that might be involved in improving systolic function or exerting CM protective effects, such as Birc6, SERCA2 and phospholamban (Fig. 3O). Together, these results reveal a distinct myocardial proteomic signature induced by GA therapy, which is associated with pro-reparative EV secretion that can recapitulate GA’s reparative effects.

### GA promotes cardiac tissue protection, angiogenesis, and reduced cardiac fibroblast proliferation

An immediate detrimental effect of tissue ischemia is loss of CMs due to necrotic and apoptotic cell death. Apoptotic cell death starts already 2h after injury and can last for weeks ^41^. Based on the proteomic data demonstrating enrichment in anti-apoptotic pathways (Fig. 3N,O), and the consistent preserved systolic functions already at 3 dpi (Fig. 2B and Fig. 3A,B), we hypothesized that reduced levels of apoptosis would be detected after GA treatment. Indeed, TUNEL assay at 24h and 96h post-injury revealed less apoptotic CMs in the ischemic border zone of GA-treated hearts (Fig. 4A-E). Additionally, we observed an increase in the expression of the anti-apoptotic protein, Bcl-XL, in the border zone and injured area of GA-treated mice (Fig. S6A,B), indicating an immediate protective effect of GA on the injured heart. These findings were further supported using a 3D *ex vivo* organ culture (EVOC) model of adult mouse hearts (Fig. S6D-H), where we exposed cardiac slices to H_2_O_2_, a reactive oxygen species (ROS) that causes cellular damage, thereby mimicking the pathophysiology of reperfusion injury. TUNEL assay and IF of γH2AX revealed reduced levels of DNA double-strand breaks in GA treated samples (Fig. S6D-H), suggesting a protective effect of GA also in the *ex vivo* model.

**Figure 4:**
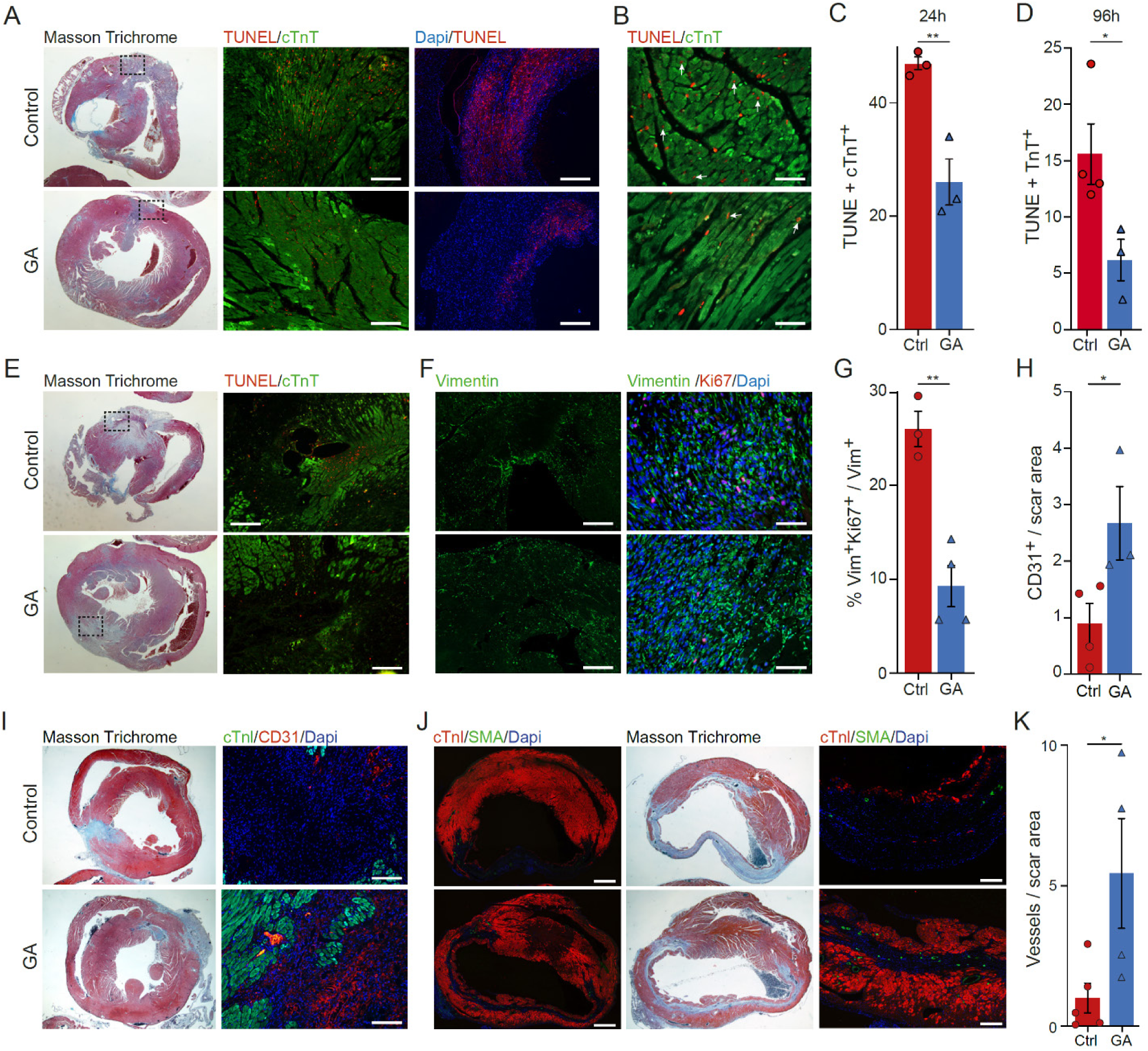
Glatiramer acetate exerts pleiotropic beneficial effect on the injured heart. **A,** Representative heart sections showing scar analysis (left) and TUNEL assay in the border-zone 24h after injury. Scale bars: middle panels, 200μm; right panels, 400μm. **B,** Higher magnifications showing TUNEL positive cells in CMs. Scale bar: 75μm. **C,D,** Quantification of percentage of apoptotic nuclei in CMs at 24h (C) and 96h (D) post-MI (n_control_ =4; n_GA_ = 3, an average of 458 cells were counted for each group). **E,** Representative heart sections showing scar analysis (left) and TUNEL assay in the border-zone 96h after injury. Scale bars: 200μm. **F,** Representative heart sections derived from control and GA-treated hearts, stained with anti-vimentin (24h) or anti-vimentin with anti-Ki67 (96h) in the border-zone. Scale bars: left, 400 μm; right, 120 μm. **G,** Quantification of proliferating CFs at 4dpi (n_GA_ = 4, n_control_ = 3). 1658 and 736 cells were counted for GA and PBS, respectively. **H,** Quantification of capillaries observed in the injured area, normalized to the scar area (n_PBS_ = 5, n_GA_ = 4). **I,** Representative heart sections derived from GA- or PBS-treated mice at 14 dpi, showing capillary formation in the scar area. Right panels show higher magnification of the left panels. Scale bars: left panels, 750 μm; right panels, 150 μm. **J,** Representative heart sections derived from GA- or PBS-treated mice at 14 dpi, showing scar area (left panels), and SMA+ blood vessels (right panels). Scale bars: 120μm. **K,** Quantification of blood vessels observed in the injured area, normalized to the scar area (n=3 for each group).

While early cardiac fibroblast (CF) activation is vital for replacing the scaffold of necrotic tissue, late and excessive CF proliferation can impair proper heart function due to stiffening of the myocardium ^42^. Staining heart sections with vimentin and Ki67 revealed intensive migration of CFs to the injured site in both GA-treated and control hearts 24h after injury (Fig. 4F). Yet, at 96h post-MI, GA suppressed CF proliferation within the scar region, relative to control (Fig. 4F,G).

Following the observed protective effects of GA and the induction of reparative inflammation, we next addressed its capacity to induce repair processes. A crucial aspect of the repair process after MI is the restoration of blood supply to the ischemic myocardium. Adequate angiogenesis can improve myocardial perfusion as well as crosstalk between CMs and endothelial cells (ECs), which improves CM function through angiocrine signals ^43^. Quantification of CD31^+^ cells revealed enhanced capillary formation within the scar area of GA-treated hearts (Fig. 4H,I), which was accompanied by ∼5-fold increase of blood vessels (Fig. 4E,F), at 14 dpi (Fig. 4J,K). This suggests that in addition to the protective effect of GA, which results in enhanced survival of blood vessels at the injured site, it can induce endothelial cell proliferation.

Taken together, these results demonstrate that GA treatment exerts pleiotropic beneficial effects that include CM protection, modulation of fibroblast activation, induction of a reparative inflammatory response and enriched vascularization, leading to improved cardiac repair and function following MI (Fig. S7).

### GA confers protection of cultured cardiomyocytes from stress-induced cell death in a cell non-autonomous manner

While the neuroprotective effects of GA are mainly attributed to its immunomodulatory properties, *in vitro* data suggest that it has a direct protective effect on cultured neurons in the absence of immune cells ^44^. To determine whether GA exerts a protective effect on CMs independent of immune cells, we used an *in vitro* cardiac culture model. Flow cytometry verified that the cultures contained negligible amounts of immune cells (Fig. S8A). Cardiac cultures derived from 3 days old (P3) mice were treated with the pro-apoptotic agent staurosporine (STS) or with H_2_O_2_. Cell death and apoptosis were analyzed using DAPI exclusion or TUNEL assays, respectively, with or without GA. CMs that were treated with STS for 18 hours in the presence of GA displayed reduced apoptosis and cell death (Fig. 5A and Fig. S8B). Control CMs under STS-induced stress lost their typical morphology and intact sarcomeric structure, whereas most GA-treated CMs maintained a typical appearance (Fig. 5A). Additionally, 48h after the addition of STS, more CMs survived in GA-treated cultures compared to control (Fig. 5B). Similarly, exposure of cardiac cultures to H_2_O_2_ revealed a significant protection of CMs in GA-treated cultures compared to control (Fig. 5C).

**Figure 5:**
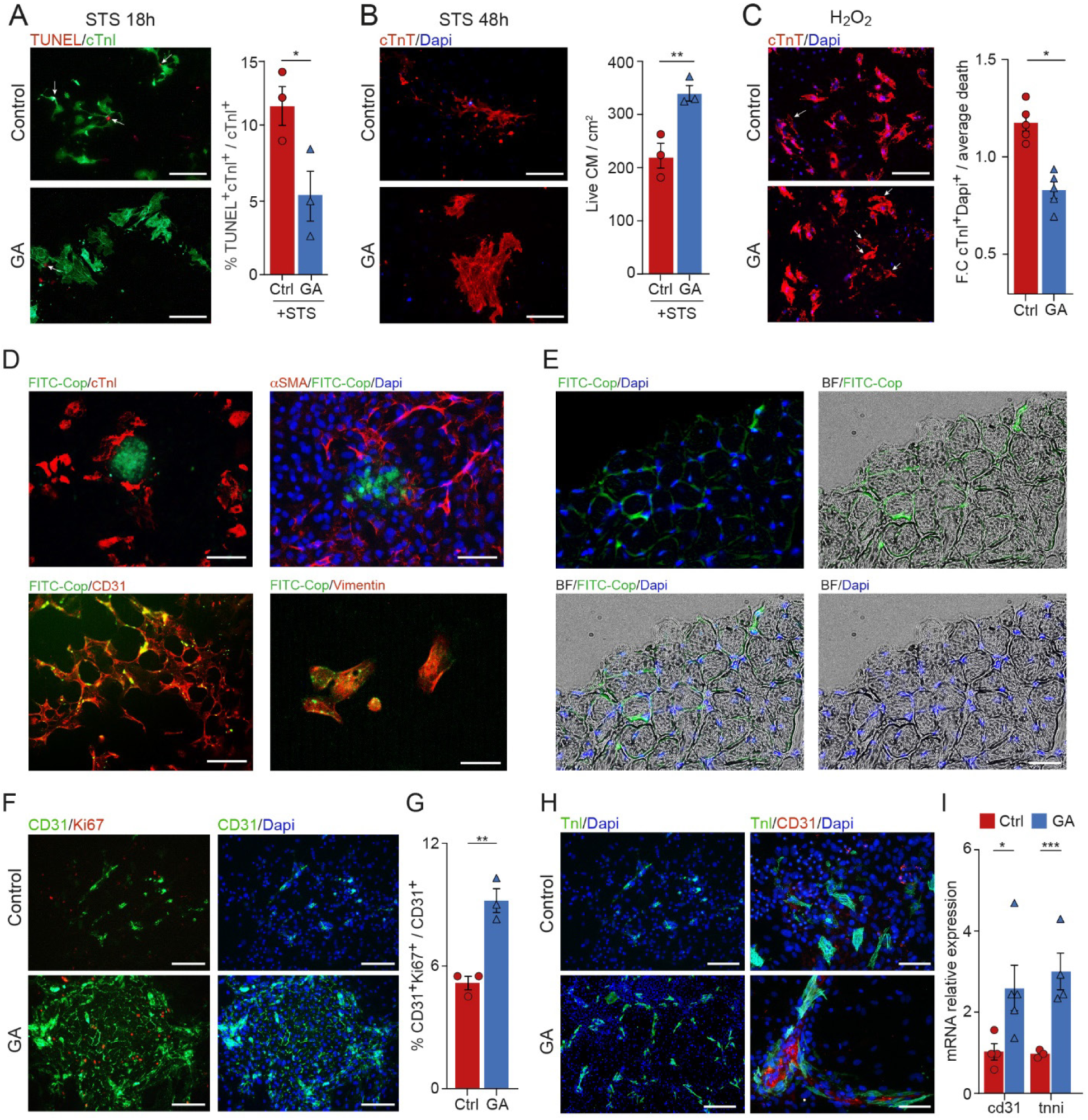
Glatiramer acetate has beneficial effects on cultured cardiac cells. **A,** TUNEL assay in P3 cardiac cultures treated either with STS alone or with STS and GA. (n=3, PBS = 4,700, GA = 1321 cells). Scale bars: 150 μm. **B,** DAPI exclusion assay at 48h after treatment. Quantification of surviving CMs per area square (right). Scale bars: 150 μm. **C,** DAPI exclusion assay after challenge with H_2_O_2_. Quantification of the fold-change in cTnI+DAPI+ dead CMs between the two groups (right) (n = 5, 2500 cells). Scale bars: 300 μm. **D,** P3 cultures were treated with FITC-GA stained with the indicated antibodies. Scale bars: upper left, 300 μm; lower left and right panels, 150 μm. **E,** EVOC treated with FITC-GA show its accumulation in interstitial cells. **F,** Representative fields show a large colony of ECs in GA-treated cultures, whereas in control cultures CD31+ cells were dispersed. Scale bars: 300 μm. **G,** Quantification of proliferating ECs (n=3 for each group, 678 and 1704 cells for the control and GA, respectively). **H,** IF stain of P3 cultures 14 days following treatment with GA. Scale bars: left, 300 μm; right, 150 μm. **I,** qRT-PCR analysis of CD31 and cTnI in the cultures described in H.

Cultured cardiac cells derived from P3 mice contain various cell types, including CMs, CFs and ECs. To detect the localization of GA in these cells, we administered GA conjugated to FITC (FITC-GA) and then labeled specific cell types using immunofluorescence staining. FITC-GA maintained its activity and induced a similar protective effect *in vitro* as unlabeled GA (Fig. S8C). Accumulation of FITC-GA started rapidly after its administration (Fig. 5D). As previously described, GA was localized in the cytoplasm of labeled cells ^44^, most of them positive for CD31 or vimentin (Fig. 5D). Interestingly, while CMs were not labeled even after prolonged incubation of 48h, they localized around the FITC-GA-labeled cell clusters. Labeling was also quite negligible in myofibroblasts, as shown by lack of FITC-GA in cells expressing SMA (Fig. 5D). The EVOC model of adult hearts showed a similar pattern of FITC-GA labeled cells, which accumulated mainly in interstitial cells and not in CMs (Fig. 5E), further supporting a paracrine effect exerted by neighboring stromal cells on the CMs.

The *in vivo* data indicating involvement of GA-derived EVs in mediating GA’s therapeutic effect, together with the absence of FITC-GA from CMs, strongly suggest a cell non-autonomous signaling as the mechanism of action whereby GA induces CM protection. To test this hypothesis, we used conditioned media from CM-depleted cardiac cultures (Fig. S8D), that were treated with either GA or control. After washing the residual GA, conditioned media was collected and used for ROS assay, i.e., challenge with H_2_O_2_ (Fig. S8E). GA-conditioned media from CM-depleted cultures reduced CM mortality following ROS exposure compared to control conditioned media (Fig. S8F), further verifying that GA acts in a paracrine manner via fibroblasts and ECs.

### GA induces spatial organization of cardiomyocytes and endothelial cells, while inhibits mouse and human CF proliferation

Next, we examined the effect of GA treatment on the growth of distinct cardiac cell populations *in vitro*. While we detected no effect on CM proliferation (not shown), GA treatment promoted the accumulation of ECs (Fig. 5F,G), further corroborating the findings obtained from the scRNA-seq analysis and the enhanced vascularization observed *in vivo*. To determine the long-term effects of GA on cardiac cultures, we followed the fate of CMs and ECs for up to 14 days of culture in the presence of GA. Immunostaining revealed a striking difference in the spatial organization of CMs. While in the absence of GA, CMs were randomly dispersed as usually observed in cardiac cultures (Fig. 5H, left panels), in GA-treated cultures, CMs rearranged in a process resembling tube formation. Co-staining revealed that GA induced tight association of CD31+ ECs with CMs (Fig. 5H, right panels). This spatial organization of ECs and CMs was previously shown to promote improved survival and beating of CMs ^45^, as well as cardiac remodeling and regeneration ^43^. RT-qPCR analysis verified the enrichment of both CM and EC populations in prolonged GA-treated cultures (Fig. 5I). Thus, GA triggers spatial association between CMs and ECs, leading to their enhanced survival. In accordance with the *in vivo* data, GA inhibited the proliferation of cultured CFs derived from either mouse hearts (Fig. S8G,H) or human cardiac biopsies (Fig. S8I).

Taken together, our *in vitro* data correlate with the observed effects of GA *in vivo* and *ex vivo*, showing that in addition to its pro-reparative immunomodulatory activity, GA directly promotes beneficial effects *in vitro* by improving CM survival, enhancing angiogenesis, and reducing CF proliferation.

### Transient treatment with GA improves heart function in a rat model of heart failure

Despite dramatic improvements in interventional cardiology that, over the past three decades, have reduced the mortality associated with acute MI, the prevalence of HF is on the rise. HF is associated with chronic inflammation. Our results in the acute MI mouse model prompted us to determine whether treatment with GA could have beneficial effects on the failing heart. For that, we induced MI in a rat model of permanent LAD ligation, allowing post-MI ventricular remodeling to progress for 28 days before starting treatment with GA or a control solvent. The severity of the injury was determined by monitoring EF values three weeks post-MI, after which the rats were divided into two experimental groups with similar measurements (Fig. 6A). The development of HF was validated by detection of ventricular remodeling by echocardiography (based on LV internal diameter in diastole) and upregulation of *nppb* in cardiac tissue (Fig. 6B,C). To prime GA-induced immunomodulation, rats were first treated with i.p GA injections daily for 7 days, followed by three weekly injections for 2 months. Echocardiography measurements were performed at the end of treatment (93 days) and 1 month later (120 days). After 2 months of treatment, the GA group displayed a significant improvement in cardiac function, as shown by a mean increase of ∼30% in EF and ∼59% in FS parameters, compared to the control group (Fig. 6D-F). Remarkably, 1 month after discontinuation of GA therapy, further improvement was observed in EF (48% increase), and the improvement in FS values was maintained (45% increase), in the GA treated group (Fig. 6E,F, and https://youtu.be/xKbnnBUaKF4, https://youtu.be/Zlu-gET60e4). Examination of individual rats revealed that most of GA-treated animals had improved EF and FS values, whereas control rats displayed worsened parameters (Fig. 6G,H). In addition to improvement in systolic function, GA-treated rats demonstrated reduced adverse remodeling of the LV, reflected by the reduced LV dimensions (Fig. 6I,J). While the control group demonstrated a persistent increase in interstitial fibrosis from 21 to 120 dpi, GA significantly inhibited this increased collagen deposition as shown by Masson Trichrome staining (Fig. 6K). These results suggest that a short 2-month GA treatment regimen results in persistent beneficial remodeling effects on the LV, reduced myocardial fibrosis and enhanced LV contractility.

**Figure 6:**
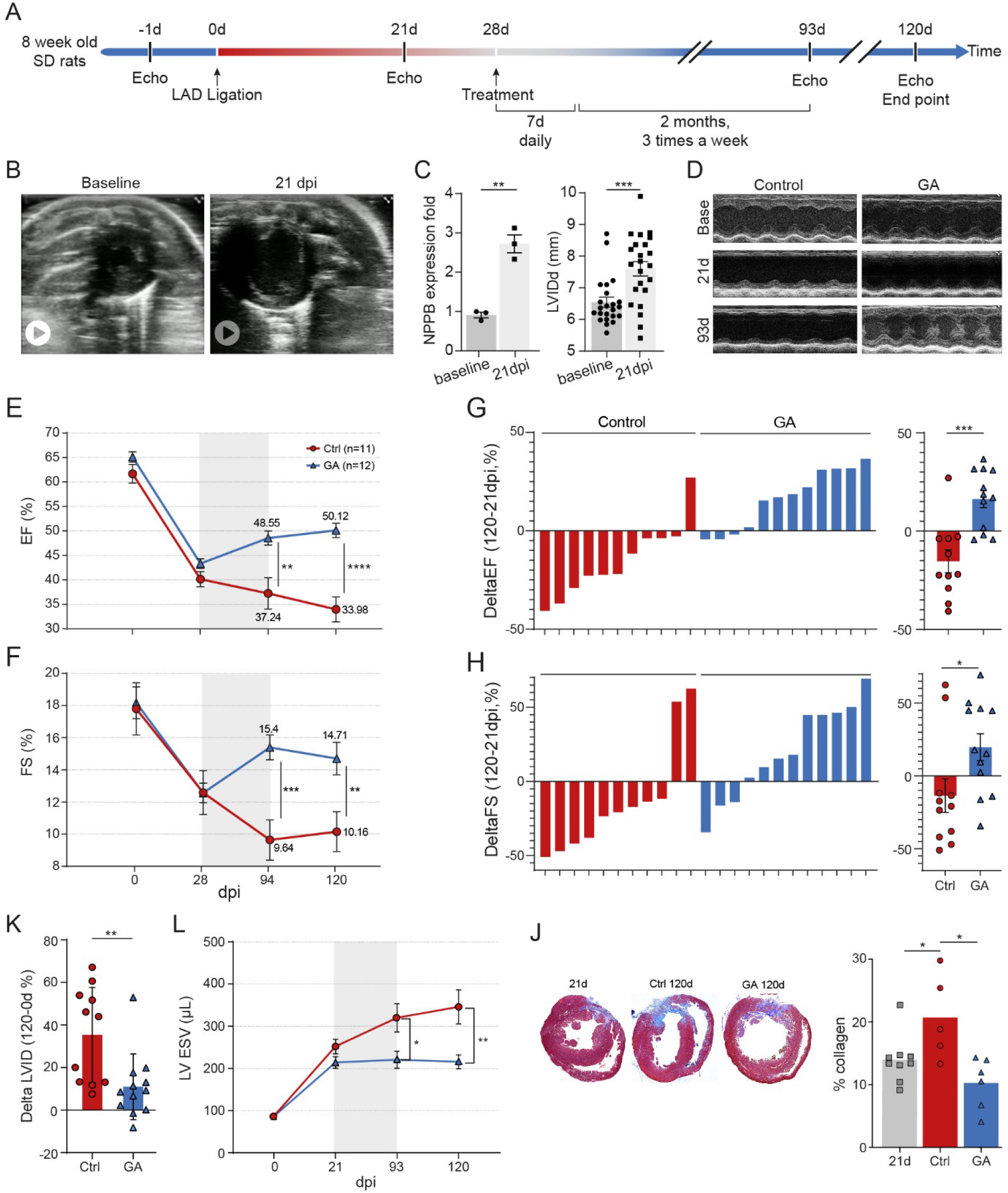
Transient treatment with glatiramer acetate improves heart function in a rat model of heart failure. **A,** A scheme of the experimental procedure. **B,** Representative echocardiographic loops of short axis view at baseline (left, https://youtu.be/k58RMQJ1ed4 and 21 dpi (right, https://youtu.be/6m3PJscTPJc) **C,** Relative mRNA expression of *Nppb* (n=3, left panel), and echocardiographic measurement of LVID in diastole at baseline and 21 dpi (n=23, right panel). **D,** Representative images using the M-Mode configuration. **E,F,** Echocardiographic measurements of EF (E) and FS (F) parameters in hearts that were treated with GA compared to control hearts. Grey area marks the period of injections. **G,H,** Graphs showing the individual (left) and mean (right) differences in EF (G) and FS (H) parameters between 21- and 120-days post-injury. **I,** The mean % difference in LVID in diastole at 120 dpi between animals treated with control (left, red) and GA (right, blue) (n_control_ = 11, n_GA_ = 12). **J,** Echocardiographic measurements of the LV volume in systole (n_control_ = 11, n_GA_ = 12). **K,** representative images of histological sections comparing interstitial fibrosis at the beginning of treatment (21d) and at end-point (120d), together with the quantification (right) (n_21d_ = 9, n_control_ = 5, n_GA_ = 6).

### GA treatment blunts the cytokine surge associated with ADHF and reduces serum levels of NT-proBNP in a small phase 2a clinical study

The clinical syndrome of HF involves a vicious cycle where myocardial and systemic inflammatory processes cause further ventricular and hemodynamic deterioration ^46^. Based on the effects we observed on myocardial injury in rodents and its excellent safety profile in humans, we postulated that GA could be repurposed to treat myocardial injury by preventing cardiac remodeling. To test this, we conducted a small proof-of-concept, phase 2a, open-label, randomized-controlled trial to assess the effects of GA treatment on the inflammatory profile of patients admitted to the hospital due to ADHF. Briefly, patients were eligible for participating if they had a confirmed diagnosis of HF with reduced systolic function (EF ≤40%), were clinically stable for the preceding month and on maximally tolerated guideline-directed medical therapy (GDMT) for at least 3 months. Major exclusion criteria included acute coronary syndrome, current infection, active treatment with anti-inflammatory agents or low compliance to medical therapy. Patients were randomly assigned in a 1:1 ratio, to receive GDMT alone or GDMT in addition to daily subcutaneous injections of GA (20mg daily), for 14 days. Baseline echocardiography was available for all patients at the time of enrollment. Patients were followed closely during hospitalization and up to 3 months thereafter. Blood samples were collected at days 0, 2, and 14 (Fig. 7A). The primary outcome was a change in serum inflammatory profile from baseline to days 2 and 14. Secondary outcomes included serum levels of cardiac biomarkers (high-sensitivity troponin and NT-proBNP), creatine kinase, and repeated hospitalizations due to ADHF within 3 months of follow-up. As this was the first clinical trial testing GA effects on patients with HF, we evaluated safety parameters, including adverse reactions, kidney function (using serum creatinine levels) and liver function (by monitoring liver enzymes).

**Figure 7:**
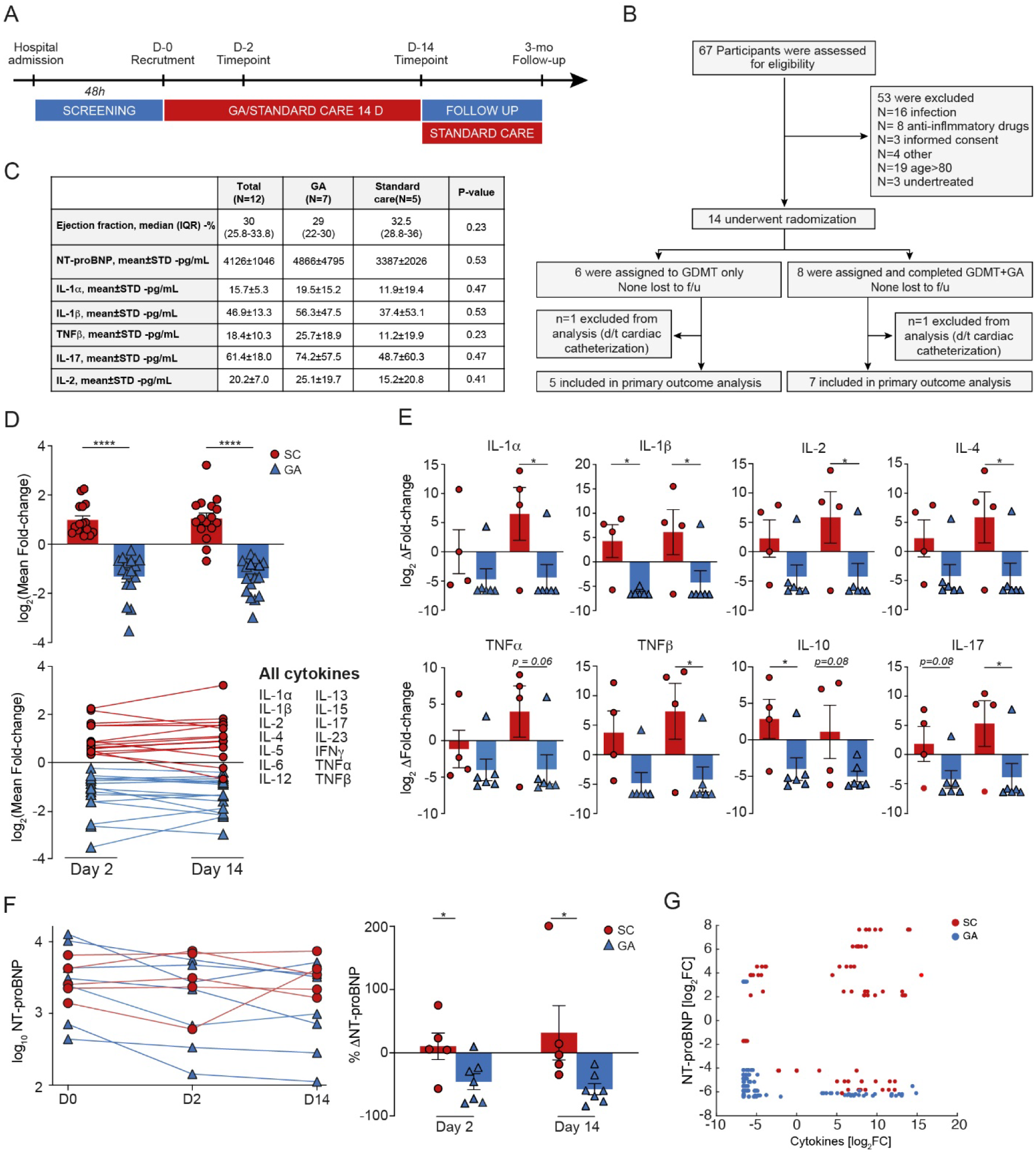
Glatiramer acetate reduces the cytokine surge and serum levels of NT-proBNP in patients with acute decompensated heart failure. **A,B,** Trial design. Patients admitted to the hospital with ADHF were screened over 48h. Eligible patients were assigned to receive only GDMT or GDMT with GA as an add-on therapy for 14 days. Repeat clinical follow up and blood sample analysis of cytokines and cardiac biomarkers were performed at the indicated time points. **C,** Baseline values of parameters associated with the primary (selected cytokines) and secondary (NT-proBNP) endpoints. Continuous variables are presented as median and interquartile range (IQR), discrete variables as proportions. **D,** Primary endpoint - GA blunts the cytokine surge in patients with ADHF. A logarithmic scale of the mean pooled results for each cytokine at days 2 and 14, normalized to baseline levels. A significant general silencing of the cytokine surge is detected in the GA-treated group of patients. **E,** Separate values of representative cytokines showing significant changes the GA-treated group. **F,** Secondary endpoint - GA treatment significantly reduced serum levels of NT-proBNP, the levels of which are presented as fold-change from baseline (left) and as raw values on a logarithmic scale (right). **G,** A scatter plot showing the fold-change in NT-proBNP levels as a function of different serum cytokines, transformed on a logarithmic scale. The GA-treated group (in blue) is clustered on the bottom left, reflecting low levels of both cytokines and NT-proBNP, as opposed to control group (in red), clustered on the top right, reflecting high inflammation and NT-proBNP levels.

Between June 2021 and June 2022, 67 patients admitted to the participating medical center with ADHF were screened. Of them, 14 patients who met the inclusion criteria were recruited. Eight patients were randomized to the GA plus GDMT group, and 6 to the GDMT alone group (Fig. 7B). Two patients (one from each group) that required in-patient cardiac catheterization were excluded from the analysis of primary outcome. There were no significant differences in baseline characteristics between the two groups (Fig. S9A and Fig. 7C). GA therapy resulted in a significant blunting of the cytokine surge associated with ADHF starting early at day 2 and maintained at day 14 following hospital admission (Fig. 7D). While the levels of serum cytokines in the standard care group increased from baseline measurements to day 14, GA-treated patients had significantly lower cytokine levels both at days 2 and 14. A post hoc analysis of individual cytokines performed after exclusion of 2 patients with abnormally high cytokine levels at day 14 (1 from each group), demonstrated a consistent trend of GA-mediated reduction throughout the various types of cytokines analyzed (Fig. 7E).

NT-proBNP is a natriuretic peptide that is mainly secreted by ventricular CMs in response to wall stretch associated with volume overload and elevated filling pressures, which are the *sine qua non* of ADHF. This cardiac biomarker serves for diagnostic as well prognostic purposes with excellent correlation with HF severity and response to therapy ^47^. Therefore, we examined changes in NT-proBNP circulatory levels in these ADHF patients with and without GA therapy. GA treatment resulted in a significant reduction in serum levels of NT-proBNP at days 2 (45%) and 14 (57%) after normalization to baseline levels (Fig. 7F, left), whereas in the control group, NT-proBNP levels increased by 10% and 31%, respectively (Fig. 7F, *p*=0.03 for both time points). When comparing the change in NT-proBNP values between baseline and day 14, a clear decrease in NT-proBNP levels was observed in the GA-treated group (−59%), while NT-proBNP levels were increased in the control group (+112%) (Fig. 7F, right and Fig. S9B). Scatter plot of cytokines and NT-proBNP levels revealed two distinct clusters of treated versus untreated patients (Fig. 7G). As an exploratory clinical outcome, we recorded the rate of re-hospitalizations due to HF during the first 3 months from the index hospitalization. Three patients (25%) were re-admitted to the hospital due to HF exacerbation (1/7 in the GA group and 2/5 in the control group) (Fig. S9C), thus the study was underpowered to show statistically significant differences. We did not detect significant differences in plasma levels of myocardial injury markers (high-sensitivity troponin or creatine kinase, data not shown). There were no mortality events observed during the follow-up period. No adverse reactions were reported throughout the course of the study, and no changes in complete blood counts and kidney or liver function were noted.

Lastly, we performed proteomic analysis of EV-enriched sera from all patients at baseline, day 2 and day 14 (Fig. S10A-C). While baseline PCA did not show separate clustering of the standard care and GA groups, a separate cluster of GA-treated patients was evident at days 2 and 14 (Fig. S10D). An unsupervised heat map generated from differentially expressed (DE) proteins at day 2 revealed a clear proteomic signature of the GA-treated patients (Fig. S10E). The main DE proteins were matricellular proteins (Tenascin, SPARCL1) that are associated with extracellular matrix remodeling, as well as proteins related to tissue stress (PRDX2 and KDM4C) (Fig. S10F). These data, obtained by an unbiased, high-throughput assay, further demonstrate the broad impact of GA treatment.

## DISCUSSION

We show here for the first time that the immunomodulator drug GA can induce reparative processes in rodent models of acute MI and ischemic HF and to significantly suppress the cytokine surge associated with ADHF in hospitalized patients. Our study is thus unique in combining basic preclinical research along with a small clinical trial suggesting GA as a potential new therapy for heart diseases. This study also stresses the potential of drug repurposing strategy, which markedly accelerates the process of drug development.

Acute MI triggers intense activation of the immune system that is required for the repair process, promoting clearance of necrotic debris, and initiating a pro-regenerative tissue response ^48^. The various stages and components of immune cell activation must be strictly regulated to prevent secondary myocardial damage, adverse remodeling, and the development of HF. As GA is primarily known for its immune modulating effects, we initially performed a through characterization of the cardiac inflammatory response following GA treatment, revealing multiple beneficial effects on neutrophils, macrophages, and T cells. These effects encompassed extensive changes in gene expression profiles, evaluated at the single cell level, as well as in immune cell tissue infiltration and cytokine secretion. It has been previously shown that an acute immune response can account for the beneficial effects attributed to cardiac stem cell therapy, specifically by modulating regional macrophages subtypes ^49^. The combined effects of reduced neutrophil activation together with up regulation in TIMD4+ resident cardiac macrophages and Tregs is expected to culminate in less collateral myocardial damage and a pro-reparative *milieu*. Indeed, we detected reduced CM apoptosis as early as 24h following injury, coinciding with the peak of neutrophil infiltration. In addition, we observed reduced CF activation, coronary vasculature protection and increased angiogenesis. We subsequently showed the therapeutic effects of GA in a rat model of ischemic HF, which further substantiated the broad capability of GA to prevent deterioration of the injured heart and enhance the reparative process.

Interestingly, apart from the known immune-modulating activity of GA, we provide evidence for pleiotropic effects *in vitro* in cardiac cultures lacking immune cells, which include CM protection and angiogenesis. The specific uptake of FITC-conjugated GA by CFs and ECs but not CMs, as well as the protective paracrine effects of GA-treated conditioned medium collected from CM-depleted cardiac cultures, suggest that these effects are, at least partly, mediated by secreted factors. The *in vivo* results of GA-derived cardiac EVs, which are enriched with proteins that promote anti-apoptotic effects and enzymes that participate in oxidative phosphorylation, support the paracrine effects observed *in vitro*, and suggest that these vesicles could also improve CM metabolism. Taken together, we propose that GA exerts a bidirectional effect; on one hand, it promotes CM protection that results in reduced immune activation, and, on the other hand, it attenuates the acute immune response, thereby minimizing secondary tissue damage (Fig. S7).

Inflammation contributes to the pathogenesis and progression of HF, and specifically of acute exacerbations ^46^. The magnitude of the inflammatory response in ADHF patients was found to be in correlation with the serum levels of NT-proBNP and is associated with a considerable risk of death within the subsequent 12 months from hospitalization ^50,51^. Therefore, we decided to test the efficacy of GA in inhibiting the cytokine surge associated with ADHF, thus potentially improving cardiovascular outcomes in this patient subset. It is important to note that the objective of this clinical study was to provide preliminary human data on the safety and potential efficacy of GA therapy to reduce the cytokine surge, neurohormonal activation and myocardial hemodynamic stress among patients with ADHF. The efficacy and high safety profile of GA have been documented in many clinical trials in large cohorts of patients treated for extended time periods ^52–54^. However, the drug has not been previously tested in patients with a heart disease, and there is a previous theoretical assumption that GA may be responsible for modulating the risk of coronary artery disease ^55^. Importantly, we did not observe any adverse effects in the treated patients, though larger patient cohorts will be necessary to confirm the safety of GA for this indication. Consistent with the results obtained in our preclinical models, we noted a profound general reduction in the levels of both pro- and anti-inflammatory cytokines after a short-term treatment with GA. The general silencing of both pro- and anti-inflammatory mediators promoted by GA is consistent with previous reports, demonstrating worse outcomes in patients in whom the general cytokine surge was not decayed ^50,51^. In addition, we detected significant reductions in the levels of NT-proBNP in the GA-treated group. As hospitalized patients with ADHF whose NT-proBNP levels remain high despite treatment are at substantially increased risk of death ^56^, GA’s effects on natriuretic peptides might be a surrogate for improved clinical outcomes.

Taken together, our findings indicate that GA may have an advantage in promoting myocardial repair over other immune-modulating agents tested thus far, as it not only targets several different aspects of the immune response and drives a pro-reparative immune phenotype, but also exerts multiple pleiotropic effects on the myocardial cellular milieu ^57^. These additive effects might be especially appealing for the treatment of HF, a disease with a considerable inflammatory component. Although exploratory in nature, the significant effects of GA detected in the small phase 2a clinical trial, and its safety use for ADHF patients, suggest a novel therapeutic potential of GA that should be assessed in larger clinical trials.

## MATERIALS AND METHODS

### Study design - animal models

#### Sample size

For the *in vivo* cardiac injury models, we used a sample size that has previously been shown to yield significant differences in cardiac parameters, such as EF, FS, and scar size. Thus, 5-10 animals were used in each experimental group, depending on the purpose of the experiment and on variation in the observed severity of injuries in individual animals. *Data inclusion/ exclusion criteria:* For the HF experiments, animals that did not show sufficient injury (EF>45%) or had too severe injury (EF <30%), as measured before the beginning of treatment, were not included in the final analysis. *Replicates:* The exact replicate number for each experiment is provided in the figure legends. *Research objectives:* The research was designed to examine the effects of GA in MI and HF rodent models and its mechanism of action. *Research subjects or units of investigation:* We used ICR female mice and Sprague Dawley rats as models of cardiac ischemic injury (acute and chronic), as well as primary mouse cardiac cell cultures. *Randomization:* In the HF experiment, animals were paired according to their EF data at 21 days post-LAD ligation and assigned randomly to either GA or control group. *Blinding:* Animals were assigned numbers and echo and histology outcomes were assessed by a blinded investigator.

### Myocardial infarction

All animal experiments were approved by the Weizmann Institute of Science Institutional Animal Care and Use Committee (IACUC) and were conducted in compliance with the guidelines and regulations regarding animal research.

#### Mice

12-week-old ICR females were sedated with 3% isoflurane (Abbot Laboratories) and then artificially ventilated following tracheal intubation. Experimental myocardial infarction was induced by permanent ligation of the left anterior descending coronary artery as previously described ^58^. Following thoracic wall closure, the mice were injected subcutaneously with buprenorphine (0.066 mg/kg^-1^) as an analgesic, and warmed until recovery. *Rats:* 8 weeks old Sprague Dawley females were anesthetized with 3% isoflurane, intubated and artificially ventilated. Buprenorphine (0.15 mg/kg SC) was administered before the surgery. Permanent LAD ligation was performed according to the previously published protocol ^59^. The rats were then warmed until recovery.

### Echocardiography parameters

Cardiac function was evaluated by transthoracic echocardiography performed on mice sedated with 2% isoflurane (Abbot Laboratories), using Vevo3100 (VisualSonics). Analysis was performed with the Vevo Lab 3.2.6 software (VisualSonics). Heart rate was continuously monitored using an ECG monitor and remained within 350-400 beats/min for the rats and 450-500 beats/min for the mice. EF and FS parameters were measured using the parasternal long axis view (PSLAX), and LVIDd was measured using the M-Mode on the parasternal short axis view (SAX).

### EV isolation and verification

Mice were subjected to permanent LAD ligation and were treated with daily intraperitoneal injections of either GA or mannitol. Animals were sacrificed at 4 dpi, the hearts were perfused with ice-cold PBS, and the LV was isolated and transferred to an Eppendorf tube with 500 µl of sterile cold PBS. The tissue was minced for 30 sec using fine scissors on ice and then centrifuged twice at 400 g for 15 min. The supernatant was centrifuged a third time at 20500 g for 45 min. Thereafter, the supernatant was transferred to 1.0 mL PC tubes (Levant Technologies, Cat., 45237) and centrifuged at 100,000 g for 1.5 h at 4°C using Sorvall MX120 Ultracentrifuge, rotor S120-AT2, fixed angle, and resuspended in 60 µl of PBS. EVs were verified according to the position statement of the International Society for Extracellular Vesicles, 2018 ^60^. Briefly, a homogenous solution of nanoparticles measuring 30-120 nm in diameter was confirmed by nanoparticle tracking analysis using NanoSight (Malvern Panalytical). Subsequently, samples were deposited on glow-discharged formvar-coated copper grids for transmission electron microscopy (TEM) and stained with 2% Uranyl Acetate. Images were taken on a Tecnai 12 TEM (Thermo Fisher Scientific), using a TVIPS F416 CMOS camera. Lastly, proteomic analysis showed enrichment of specific EV markers.

### EV-mediated reduction in scar area

Mice were subjected to permanent ligation of LAD and received daily treatment with either 2mg GA, i.p, or an appropriate solvent for 3 days. Subsequently, EVs were isolated from the LV, and used to treat a second cohort of mice using a single intramyocardial (i.m.) injection following LAD ligation. We validated that GA-derived EVs and control-derived EVs were at a similar concentration using NTA (approximately 4 × 10^9^ particles/mL) in a volume of 50 uL.

### EV proteomic analysis

EVs were isolated according to protocol. After ultracentrifugation, pellets were kept at −80°C until analysis. The samples were lysed and digested with trypsin using the S-trap method. The resulting peptides were analyzed using nanoflow liquid chromatography (nanoAcquity) coupled to high resolution, high mass accuracy mass spectrometry (QE HF). Each sample was analyzed separately in a random order in discovery mode. The raw mass spectrometry data were processed with MetaMorpheus version 0.3.20. Data were searched against the mouse UniproKB XML database version 01_2022, with common lab proteins. We applied the spectral recalibration module, G-PTMD module and Search module. Data were normalized and filtered for maximum 1% FDR. Resulting protein table was imported to Perseus. Data were filtered for replication in at least 70% of the replicates in at least one group. Log transformed and missing values were imputed from a low, random distribution. ANOVA was used for statistical evaluation. We used the following threshold for significance: q-value <0.05, fold-change >2, and at least 2 peptides detected. Gene ontology and pathway analysis was performed using DAVID Analysis Tool, ranked according to log10(control-p-value).

### Cell culture

Primary cardiac cultures were isolated from P3 mice using a neonatal dissociation kit (Miltenyi Biotec,130-098-373) and the gentleMACS homogenizer, according to the manufacturer’s instructions. Cells were cultured in DMEM/F12 medium (Sigma, D6421) as previously described ^58^. The medium was replaced every other day. For most experiments, cells were seeded in 96-well plates at a cell density of 20,000-30,000 cells/well, or at a density of 2-2.5 x 10^5^/6 well. Treatment with GA (20-30 μg/ml) or control started 4 days after isolation. Cells from FUCCI mice (a generous gift from Dr. Mark Sussman, San Diego State University, CA, USA ^61^) were used to follow CM cell cycle.

### Clinical trial

The study was conducted according to the guidelines of the Declaration of Helsinki and approved by the Institutional Review Boards of Hadassah University Medical Center and the Israeli ministry of health (approval number: HMO-093420), and is registered in Israel Ministry of Health clinical trials site (https://my.health.gov.il/CliniTrials/Pages/MOH_2021-05-10_009957.aspx). The study was performed in Hadassah Medical Center, Jerusalem, Israel. Basic laboratory work (complete blood count, serum electrolytes, creatinine, NT-proBNP, hs-troponin, CK and liver enzymes) were performed by the core laboratory of the Hadassah Medical Center as part of the routine clinical patient care. Immune panels, consisting of pro- and anti-inflammatory cytokines, were performed in an independent core research laboratory in Sheba Medical Center, Israel. All laboratory work-up was performed by an independent core laboratory that was blind to patient assignment. All patient samples were anonymous and coded. Cytokine levels were measured using Q-Plex^TM^ Human Cytokine Screen (16-Plex), Q110933HU, and Q-Plex^TM^ Human Chemokine (9-Plex), Q120233HU, Quansys Biosciences, UT, USA.

### Statistical analysis

The results are presented as means ± SEM and the number of independent biological repeats is indicated for each experiment. Statistical comparisons were carried out by two-tailed Student’s *t* test or analysis of variance as appropriate, followed by correction for multiple comparisons using Tukey or Newman-Keuls procedure. Data are presented as mean ± SEM. Statistical significance was calculated using a two-tailed *t*-test, * *P* < 0.05, ** *P* < 0.01, *** *P* < 0.001.

## Data Availability

All data produced in the present study are available upon reasonable request to the authors

## Acknowledgments

We thank Ishai Sher from the Graphic Department at Weizmann Institute for excellent graphic assistance; Marina Cohen from the Histology unit at Weizmann Institute; Yishai Levin and Corine Katina from the Proteomic Unit of the Grand Israel National Center for Personalized Medicine, at Weizmann Institute; Galina Levin, Larisa Kogan and Lilian Abu-Heichel for coordinating the clinical trial in Hadassah Medical Center; Oliana Vazhgovsky for coordinating the biopsies transfer from Sheba Medical Center.

## Funding

The study was supported by the following funding sources:

European Research Council, ERC AdG grant no. 788194, CardHeal (E.T)

ERC-PoC no. 899224, ReDHeaD (E.T)

EU Horizon 2020 Research and Innovation Programme REANIMA

## Author contributions

Conceptualization: R.S, E.T, G.A, O.A, K.U

Methodology: R.S, E.T, G.A, O.A, R.A, K.U, Ru.A, Ri.A

Investigation: G.A, J.E, K.U, H.B, R.S, S.M, D.L, D.K, U.K, D.N, M.A, D.M, T.S, L.Z, Z.P

Funding acquisition: E.T Supervision: E.T, R.S

Writing – original draft: E.T, R.S, G.A

Writing – review & editing: O.A, R.A

## Competing interests

The authors vouch no competing interests.

## Data and materials availability

Raw data of proteomic analysis will be provided at later stages.

## List of Supplementary Materials

Supplementary Materials and Methods Fig. S1 to S10

Supplementary Tables 1-2

## Supplementary Materials

### Supplementary materials and methods

#### Glatiramer acetate

Two types of GA were used for the experiments: either powder or Copaxone syringes (both from Teva), with proper controls for each type. PBS was used to dissolve the powder, whereas mannitol was the solvent of Copaxone syringes. The potency of the two GA preparations was very similar in all the described experiments. For the *in vivo* experiment, we used a well-established concentration of 2 mg/100 µl, which was shown to be effective for the treatment of EAE mice ^9,10^. The concentration used for the *in vitro* experiments was 20-30 µg/ml. A Protein FITC Labeling Kit (EZLabel^TM^, K832-5, BioVision Inc., USA) was used to produce a FITC-conjugated GA, according to the manufacturer’s instructions.

#### Scar quantification

Scarred tissue was quantified by performing Masson’s trichrome staining on serial cardiac sections spanning the entire left ventricle using ImageJ. The area of scar (blue) and LV were calculated and summed from all sections in order to obtain true scar volume normalized to LV myocardial volume.

#### Blood vessel and endothelial cell density

Capillary and blood vessel density were calculated by measuring CD31+ area and counting αSMA+ blood vessels in scar tissue, respectively, normalized to scar area. Area measurements were performed using ImageJ.

#### Assessment of ischemic area-at-risk and infarct zone

Assessment of IAAR was performed as previously published ^62^ with modifications. Briefly, At the 4^th^ day after permanent LAD ligation, animals were anesthetized using CO_2_ inhalation. Demarcation of ischemic area-at-risk was achieved by performing *in situ* antegrade perfusion using 8 mL of black dye through ventricular apical canulation. Subsequently, hearts was excised from the chest, washed twice in ice-cold PBS and transferred to −20^0^c for 8 minutes. Hearts were then axially sectioned using a razor blade, at 200 μm intervals. For delineating infarct zone, the myocardial sections were subsequently incubated at room temperature in 2% triphenyltetrazolium chloride (Sigma-Aldrich, T8877) for 10 minutes, after which the sections were washed twice with PBS. Sections were imaged using a stereomicroscope (Carl Zeiss Stemi 305cam Binocular). Quantification of ARR and IZ were performed using ImageJ, and normalized to LV where appropriate.

#### Sample and libraries preparation for scRNA-seq

Single-cell RNA-seq libraries were prepared at the Crown Genomics Institute of the Nancy and Stephen Grand Israel National Center for Personalized Medicine, Weizmann Institute of Science using the 10X Genomics technology. Tissue was dissociated into single-cell suspension using a neonatal dissociation kit (Miltenyi Biotec,130-098-373) and the gentleMACS homogenizer, according to the manufacturer’s instructions. Briefly, animals were sacrificed, and the hearts were perfused in situ using ice-cold PBS solution. Subsequently the hearts were excised from the body and washed twice in PBS. The atria and right ventricle were removed to enrich for LV tissue. The LV-enriched tissue was minced to small pieces using fine scissors. The tissue was transferred to a gentleMACS C-tube (130-093-237) and dissociation was carried according to the protocol. Enrichment for CD45+ cells was achieved using CD45 MicroBeads enrichment protocol (Miltenyi Biotec, 130-052-301). Briefly, CD45+ cells were magnetically labeled with CD45 MicroBeads. Positive selection of immune cells was achieved by mounting the labeled suspension on a MACS Column, placed in a magnetic field of a MACS Separator. Cells were counted and viability was assessed using trypan blue and a hemocytometer. Cells were diluted in PBS + 0.04% BSA to a final concentration of 1000 cells/ul and immediately processed with the Chromium Next GEM Single Cell 3’ v3.1 kit, according to the manufacturer protocol. Final libraries were quantified by qPCR with the NEB-next Library Quant Kit (New Englnad Biolabs), as well as with Qubit and TapeStation. Sequencing was done on a Nova-Seq6000 using SP, 100 cycles kit mode allocating 800M reads in total (Illumina). Fastq files were generated by the usage of bcl2fastq v2.20.0.422.

### Bioinformatic analysis

CellRanger pipeline (Zheng et al., 2017) (v7.0.1, 10× Genomics) with default settings was used for alignment (mm10 reference genome, 2020-A version, downloaded from 10× Genomics website), filtering, barcode counting and UMI (Unique Molecular Identifier) counting. The R Seurat package (Satija et al., 2015) (v4.0.4) was used for quality control, dimensionality reduction, visualization, and analysis. Low-quality cells were excluded based on the following criteria: having fewer than 250 genes, fewer than 500 UMIs, and exceeding 4 MAD (median absolute deviation) the median for genes, UMIs, and mitochondrial reads. Additionally, genes expressed in less than 10 cells were eliminated from the analysis. After implementing these quality control measures, a total of 49,369 cells (from all samples) and 21,031 genes were retained for further analysis (control dpi 1: 3016, GA dpi 1: 3537, control dpi 4: 24431, GA dpi 4: 18,385).

Merged counts matrix was log-normalized using the NormalizeData function. The 2,000 highly variable genes were identified using the FindVariableFeatures function with the ‘vst’ method. Data was scaled with the ScaleData function. Principal component analysis (PCA) was performed. Seurat’s unsupervised graph-based clustering and uniform manifold approximation (UMAP) were conducted on the projected principal component (PC) space.

Marker genes for each cluster were determined with the non-parametric Wilcoxon rank sum test by FindAllMarkers function. Those with a logFC > 0.5 and expressed in at least 50% of the cells were selected as significant marker genes. Cell types were identified based on the expression of classic marker genes. Cells were represented by a total of 19 clusters including monocytes and macrophages (*Cd68+Itgam+*), neutrophils (*S100a9+S100a8+*), T cells (*Cd3e+Cd3d+*), B cells (*Cd79aMs4a1+*), endothelial cells (*Kdr+Pecam1+*), fibroblasts (*Col1a1+Dcn+*), myofibroblasts (*Col1a1+Postn+*), pericytes (*Rgs5+Abcc9+*) and smooth muscle cells (*Acta2+Myh11+*) (Fig. 2C). Visualization used Seurat’s FeaturePlot, DotPlot and VlnPlot functions.

FindMarkers was used to identify DE genes between different clusters. Those with a p_val_adj < 0.05 were considered to be significant and taken for furher analysis using IPA (Ingenuity® Systems, www.ingenuity.com) or DAVID analysis tool.

#### Bulk proteomic analysis

Cardiac samples of proteomic analysis were processed as previously published ^58^. Hearts were removed 4 days post-MI and briefly perfused with ice-cold PBS. The left ventricle was dissected and snap-frozen in liquid nitrogen. Samples were crushed to a fine powder, intermittently maintaining nitrogen cooling, and transferred into a lysis buffer consisting of 5% SDS and 50 mM Tris (pH 7.4) for thorough homogenization using a drill. The samples were centrifuged at 5000 g for 15 sec and boiled at 96°C for 15 min. After further centrifugation at maximal speed for 2 min, the supernatant was collected, and protein concentrations were quantified by BCA assay (Thermo Fisher, 23225). For LC-MS and data processing, samples were lysed with urea and subjected to in-solution tryptic digestion, followed by a desalting step. The resulting peptides were analyzed using Waters HSS-T3 column on nanoflow liquid chromatography (nanoAcquity) coupled to high resolution, high mass accuracy mass spectrometry (Fusion Lumos). Thresholds used were p-value <0.05, fold-change >2, and at least 2 peptides detected on mass spectrometry. Gene ontology (GO) terms and pathway analysis performed using DAVID Analysis Tool.

#### ROS assay

ROS assays were performed as previously published with modifications ^63^. P3 primary cultures were treated with either 30 µg/ml GA, PBS, or conditioned media for 3 days. On the third day of treatment, the cells were challenged with 1 µM H_2_O_2_ (Sigma, 7722-84-1) or a similar volume of PBS for 2 h, after which the wells were washed twice with warm PBS. For DAPI exclusion assay, 0.5 µg/mL DAPI (Sigma, Cat. no. D9542) was added to the media for 10 min. Cells were subsequently washed with PBS and fixated in 4% PFA followed by immunofluorescence staining.

#### RT–qPCR

RNA from whole hearts or cultured cells was isolated using a NucleoSpin kit (Machery Nagel, 740955.50) according to the manufacturer’s instructions. A High capacity cDNA reverse transcription kit (Applied Biosystems, 4374966) was used to reverse transcribe 1 µg of purified RNA according to the manufacturer’s instructions. The quantitative PCR reactions were performed using a Fast SYBR Green PCR master mix (Thermo Fischer Scientific, 4385614) on a StepOnePlus Real-Time PCR system (Applied Biosystems). Values for specific genes were normalized to *Hprt* housekeeping control.

#### TUNEL assay

Apoptotic cell death of both cell cultures and cardiac sections was measured using the ApopTag® Red *In Situ* Apoptosis Detection Kit (S7165, Merck), according to the manufacturer’s instructions. Staurosporine (1 µM, Sigma-Aldrich) or DMSO were added to cardiac cultures together with either 20 µg/ml GA or PBS for 18 h.

#### Ex vivo organ cultures (EVOC)

EVOC were prepared according to Gavert *et al.,* ^64^, with modification for heart samples. Adult mice were euthanized, and their hearts were placed in ice-cold PBS. The fresh heart tissues were then filled with a 3.5% (w/v) low melting agarose preparation media sliced into 280-μm-thick slices using a vibratome (Leica VT1000 S). Heart slices were cultured in growth media with GA or Mannitol for 48h, followed by treatment with 0.1 µM H_2_O_2_ for 4h. Slices were fixed, embedded in paraffin, and 5 μm slices were used for TUNEL assay and IF staining.

#### Cytokine measurements

Plasma and tissue protein levels were measured using a multiplex ELISA kit (mouse cytokine array GSM-CYT-1, RayBiotech, GA, USA) according to the manufacturer’s protocol. In brief, blood was drawn from the retro-orbital venous plexus using glass pipettes pre-washed with 500 U/ml of heparin. Samples were centrifuged at 1200 rpm for 10 min at 4°C and plasma was transferred into new tubes and kept at −80°C. For tissue protein analysis, LV tissue was dissected from the whole heart and snap-frozen in liquid nitrogen. Frozen samples were crushed with a mortar and pestle and subsequently homogenized with a drill. Total cell lysates were isolated using RIPA buffer supplemented with 1:100 protease (Sigma, P8340) and 1:100 phosphatase inhibitor cocktails (Sigma, P5726 and P0044). Protein concentrations were measured by BCA assay (Thermo Fisher, 23225). To each well, 750 µg/mL of tissue protein extract or 1:2 diluted plasma were added and incubated overnight at 4°C with blocking solution. On the following day, the samples were washed and incubated with biotinylated antibody cocktail and a secondary Cy3 equivalent dye-streptavidin. The slides were read by the RayBiotech array scanning service and data were analyzed using Piezoarray Software, version 1.1.0.0016.

#### EV-enriched serum from patients

Enrichment for human EVs was performed from patients’ sera using similar protocols. Subsequently, the sera were analyzed using the standard methods for EV detection: NTA, EM and verification of EV-specific markers using mass spectrometry.

#### Bulk RNA-seq

For RNA-seq, GA- and control-treated hearts were collected 24 h after MI, and RNA samples were purified using the miRNeasy kit (1038703, Qiagen) according to the manufacturer’s instructions. Sequencing libraries were prepared using the RNA-seq protocol of the Nancy and Stephen Grand Israel National Center for Personalized Medicine (G-INCPM), Weizmann Institute of Science. Single-end short reads were sequenced on Illumina NextSeq machine. Sequencing libraries were constructed with barcodes to allow multiplexing of the samples, yielding a median of ∼25 million single-end reads per sample. Briefly, total RNA was fragmented followed by reverse transcription and second-strand cDNA synthesis. The double-strand cDNA was subjected to end repair, A base addition, adapter ligation and PCR amplification to create libraries. Libraries were evaluated by Qubit and TapeStation. For RNA analysis, adapters were trimmed using the cutadapt tool. Resulting reads shorter than 30 bp were discarded. Reads were mapped to the *Mus musculus* reference genome GRCm38 using STAR, supplied with gene annotations downloaded from Ensembl, with EndToEnd option and outFilterMismatchNoverLmax set to 0.04. Expression levels for each gene were quantified using htseq-count, using the gtf above. Differentially expressed genes were identified using DESeq2 with the betaPrior, cooksCutoff and independentFiltering parameters set to False. Raw p-values were adjusted for multiple testing using the Benjamini and Hochberg procedure. Pipeline was run using snakemake.

#### Human cardiac cultures

Cardiac biopsy specimens were obtained from patients with a diagnosis of tetralogy of Fallot (TOF), undergoing an open-heart surgery. Tissue explants were kept on ice until processed and were mechanically minced into 1-mm^3^ pieces. The cardiac human cells were isolated using a neonatal dissociation kit (Miltenyi Biotec,130-098-373) and the gentleMACS homogenizer, according to the manufacturer’s instructions. The study was conducted according to the guidelines of the Declaration of Helsinki and approved by the Institutional Review Boards of Sheba Medical Center.

#### Immunofluorescence staining

##### Tissue cultures

cells were fixed with 4% PFA in PBS for 10 min and permeabilized with 0.2% Triton X-100 in PBS for 5 min, followed by blocking with PBS containing 0.1% Triton X-100 and 3% bovine serum albumin for 1 h at room temperature. Immunostaining was performed as previously described ^58^. *Cardiac sections:* paraffin-embedded hearts were sectioned, deparaffinized and stained as above with the addition of an antigen retrieval step that was performed in 10 mM citric acid, (pH 6.0) or 0.1 M Tris-EDTA buffer (pH 9.0). Images were obtained with a Nikon Eclipse Ti2 fluorescent microscope with the Nikon’s NIS-Elements software. Primary antibodies: anti-pHistone 3 (pH3,ab47297 ABCAM 1:1000), anti-Ki67 (Cell Marque #275R, 1:200), anti-cTnT (Abcam, ab33589, 1:200 or CT3 hybridoma product, DSHB by Lin, J.J.-C., 1:10), anti-cTnI (Abcam, ab47003, 1:200 or TI-4 hybridoma product, DSHB by Schiaffino, S., 1:10), anti-CD31 (Abcam, ab28364, 1:200), anti-Vimentin (Abcam, ab24525, 1:200). SMA (Sigma, A2547, 1:400), anti-Foxp3 (Invitrogen, FJK-16S, 1:100), anti-CD3 (Abcam, ab16669, 1:150).

#### FACS

Hearts were perfused with 10 ml of ice-cold PBS and left ventricles were dissected and digested in RPMI containing Collagenase-I (Sigma, 450 U/ml), Collagenase-XI (Sigma, 120U/ml), Hyaluronidase (Sigma, 60 U/ml) and DNase-I (Sigma, 10 mg/ml) at 37°C for 1 h, while agitated at 200 rpm. Digested materials were passed through a 120-µm metal mesh into 12 ml PBS and pelleted by cooled centrifugation (4°C, 1400 rpm, 5 min). Supernatant was removed and the pellet was resuspended in 3 ml of MACS buffer (2% FBS, 1 mM EDTA in PBS), followed by additional centrifugation (4°C, 1400 rpm, 5 min). Samples were blocked in 50 µl of anti-CD16/32 antibodies (clone: 93, 1:200) for 20 min at 4°C, resuspended in 3 ml MACS buffer and centrifuged (4°C, 1400 rpm, 5 min). Cells were incubated in antibody mixture (50 µl) in MACS buffer for 20 min at 4°. Samples were washed in 3 ml MACS buffer containing DAPI (0.5 µg/mL) and centrifuged (4°C, 1400 rpm, 5 min). Single-cell suspension was resuspended in 0.5 ml of MACS buffer and filtered through 40-µm mesh prior to FACS analysis by Fortessa (BD Biosciences, BD Diva Software). For intracellular staining, cells were processed as described above with the following changes: after primary antibody incubation, cells were washed in MACS buffer and centrifuged (4°C, 1400 rpm, 5 min), followed by fixation using 1 mL of True-Nuclear™ (BioLegend, cat# 424401) for 18h at 4°C. Cells were processed further according to the True-Nuclear™ manufacturer protocol. Prior to FACS analysis, cells were resuspended in 0.5 ml MACS buffer. Data were analyzed using the FlowJo (v10.6.2) software. *Antibodies* (all from BioLegend): CD45 (clone: 30-F11, 1:100), CD11b (clone: M1/70, 1:200), CD4 (clone: GK1.5, 1:100), TCRb (clone: H57-597, 1:100), FOXP3 (clone: MF-14, 1:100), LY6G (clone: 1A8, 1:100), LY6C (clone: HK1.4, 1:100), GR1 (Ly6C+, Ly6G+, clone: RB6-8C5, 1:100), F4/80 (1:80), Tim-4 (clone: F31-5G3, 1:100), CCR2 (clone: SA203G11, 1:100). *Gating for FACS:* Myeloid cells: DAPI-CD45+CD11b+; neutrophils: DAPI-CD45+CD11b+Ly6g+ cells, further parsed by LY6C; T-regulatory cells: DAPI-CD45+CD11b-TCRb+CD4+, further parsed by FOXP3; resident macrophages: DAPI-CD45+CD11b+F4/80+ GR1-(Ly6c+,Ly6G+), further parsed by TIMD4+CCR2-.

#### Conditioned media

P3 primary cultures were seeded on 96-well plates (Nuncio, 167008) at a density of 50,000-60,000 cells/well. We used the pre-plating technique to exclude CMs^65^, by washing the cultures 3 h after isolation and growing only the adherent cells, which were mainly CFs, myofibroblasts and ECs. Two days afterwards, the cells were incubated with either mannitol or 30 µg/ml GA for 10 h, after which they were washed twice with PBS to discard all residual GA and incubated in fresh medium. The medium was collected after 12 h, passed through a 0.22-µm cell strainer (Durapore Membran, SLGV033R, Merck Millipore, Cork, IRL) and kept at −20°C until use for ROS assay.

## Supplementary Figures

**Figure S1:**
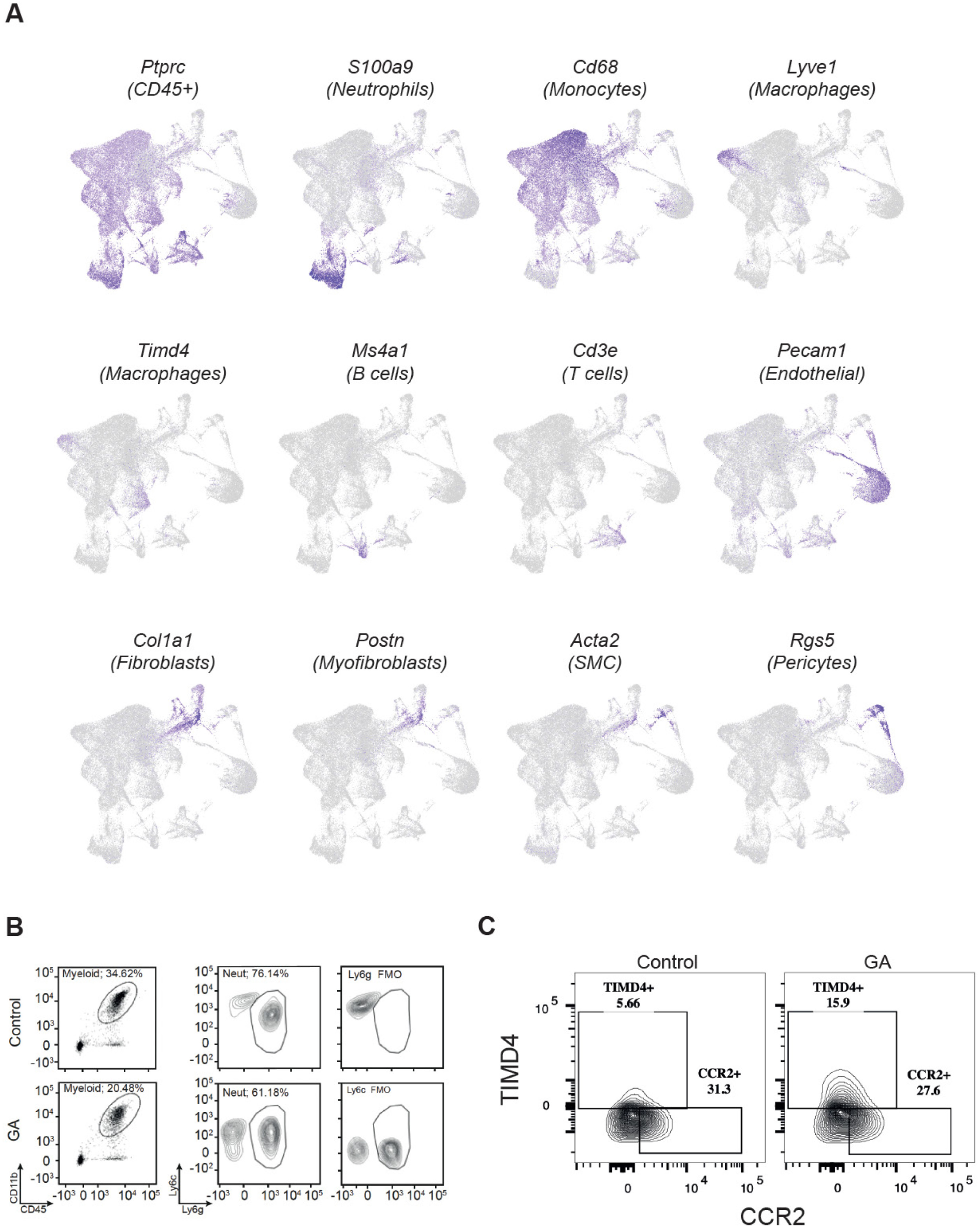
Cluster annotations and flow cytometry gating. **A**, Distinct cell populations are visualized in UMAP dimensionality reduction plots. Cluster annotation was performed using canonical markers for each cell type. **B**, Myocardial neutrophil infiltration was analyzed using FACS. Gating data are provided. **C**, number of myocardial Timd4+ resident macrophages was analyzed using FACS. Gating data are provided.

**Figure S2:**
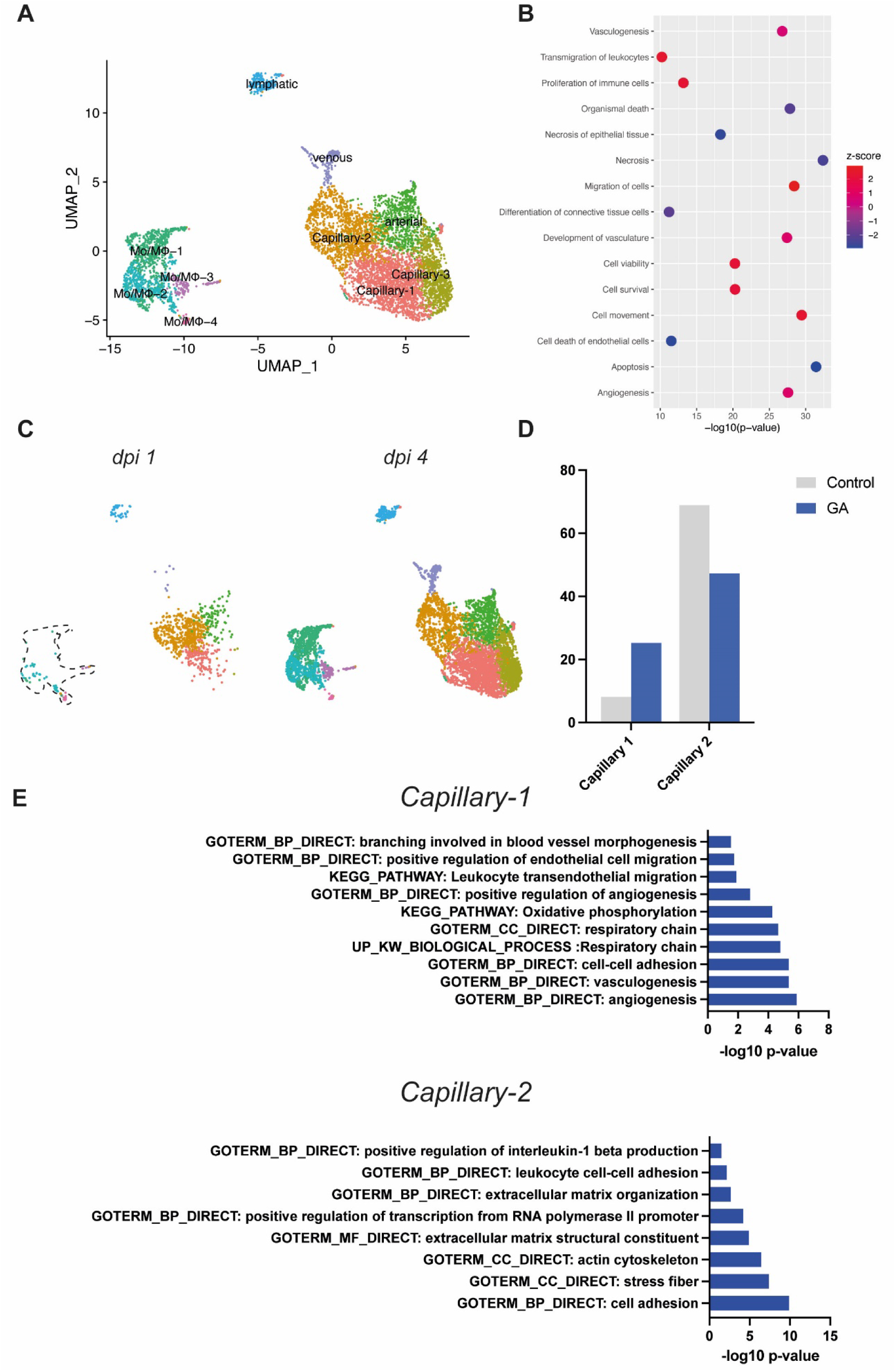
GA promotes a protective and proangiogenic effect on endothelial cells at 1 dpi. **A**, A targeted UMAP performed on the subset of endothelial cells. **B**, IPA performed on DEG comparing GA and Control on the cluster of ECs from the general UMAP (adjusted *p*-value <0.05). **C**, Capillary-3 and monocytic clusters accumulate on 4 dpi. **D**, A frequency plot, showing the relative abundance of endothelial capillaries 1 and 2 at 1 dpi. Capillary-2 was more abundant in the GA-group, while Capillary-1 was more abundant in the control group. **E**, An unbiased heatmap of the cluster markers of Capillary-1 and Capillary-2 (adjusted p-value <0.05). **G**, Pathway analysis using the DAVID tool of the cluster markers for Capillary-1 and Capillary-2.

**Figure S3:**
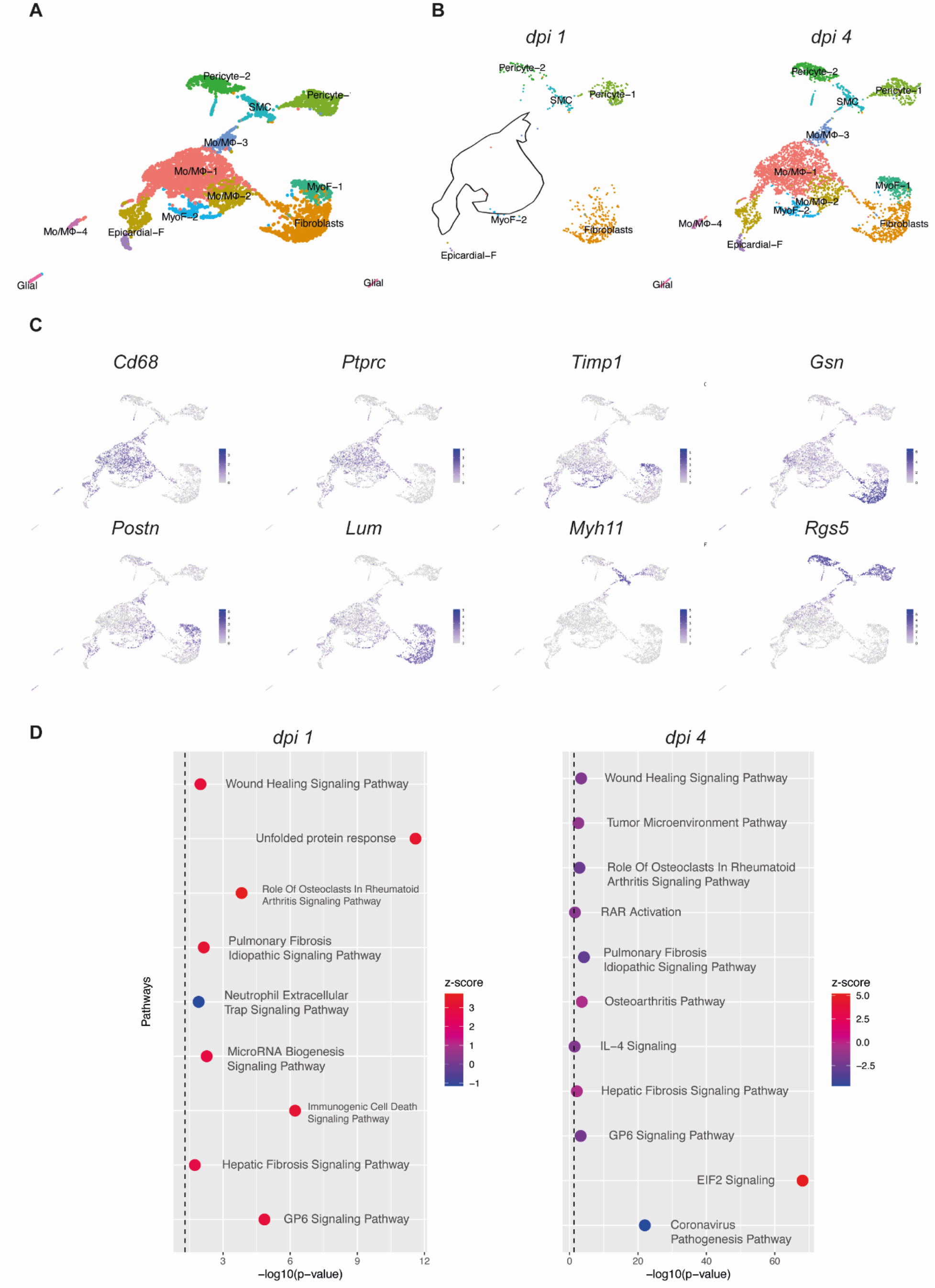
GA promotes a biphasic fibroblast response following acute MI in mice. **A,** A targeted UMAP of fibroblast subset from the general UMAP showing a distinct cluster separation of fibroblasts, double-marker monocytes positive also for fibroblast genes, probably reflecting fibroblast phagocytosis, pericytes and glial cells. **B**, Myofibroblasts as well as the double-markers monocytes are absent at 1 dpi, and they strongly appear at 4 dpi. **C**, cluster markers used for annotations. **D**, IPA performed at Days 1 and 4 post injury on the DE genes between GA and Control of the quiescent fibroblast cluster (adjusted P-value <0.05). While at 1 dpi there is a strong fibroblast activation, an important contributor for wound healing, at 4 dpi there is a marked fibroblast silencing, that might reflect a decreased tendency for a fibrotic scar formation.

**Figure S4:**
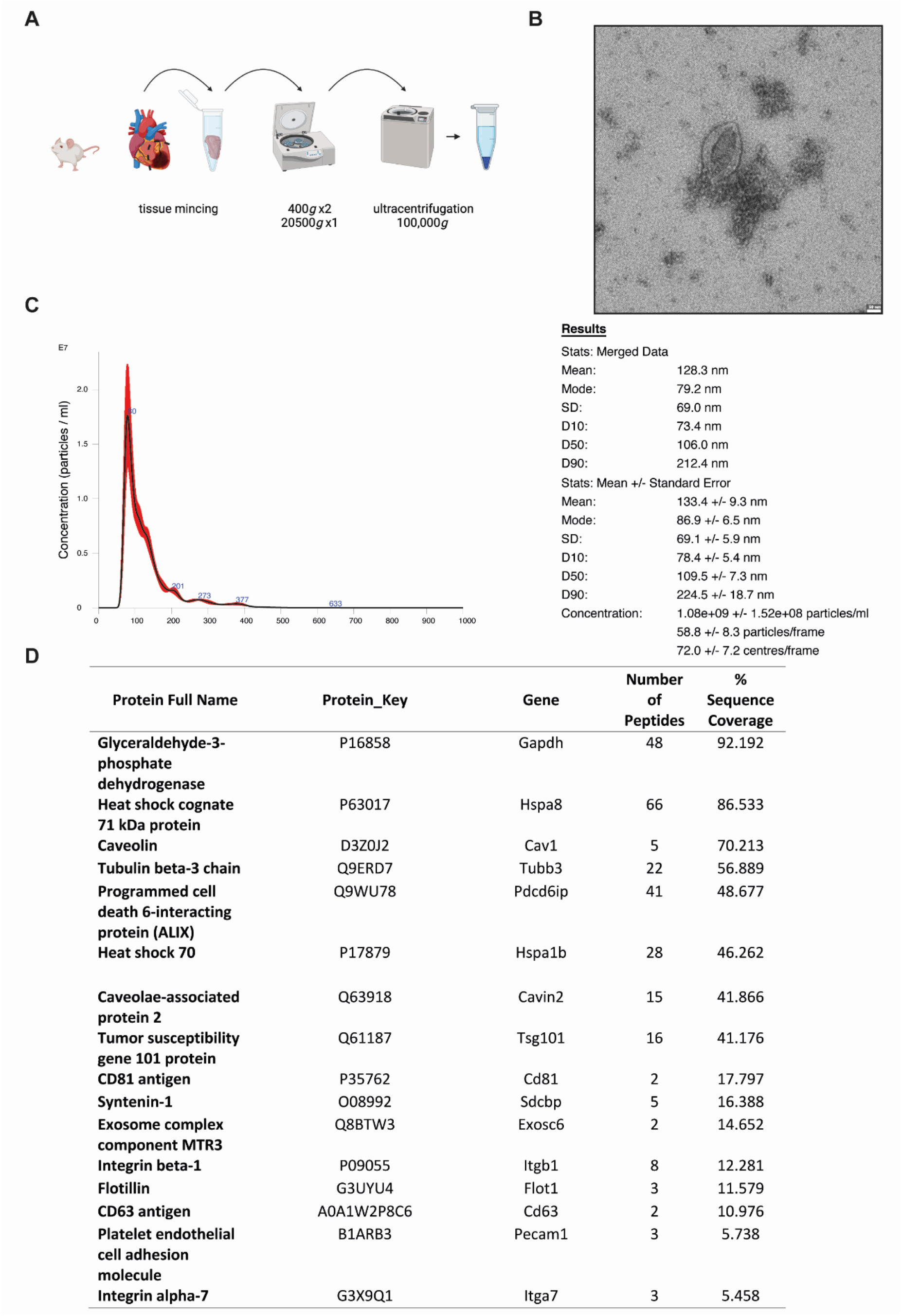
Validations for EV isolation. **A**, Schematic illustration for EV isolation protocol from mouse LV tissue. EVs were purified by sequential centrifugations at increasing speed up to 100,000 *g*. Subsequently, EVs were analyzed to confirm high-purity isolation. **B,** Transmission electron microscopy. Samples were stained with 2% uranyl acetate, revealing membrane-enclosed particles of the anticipated diameter. Scale: 100 nm. **C**, Nanoparticle tracking analysis (NTA) demonstrates a unimodal pick below 200 nm, consistent with EVs diameter. **D**, Data from proteomic analysis. Common EV-specific protein markers were detected using mass-spectrometry.

**Figure S5:**
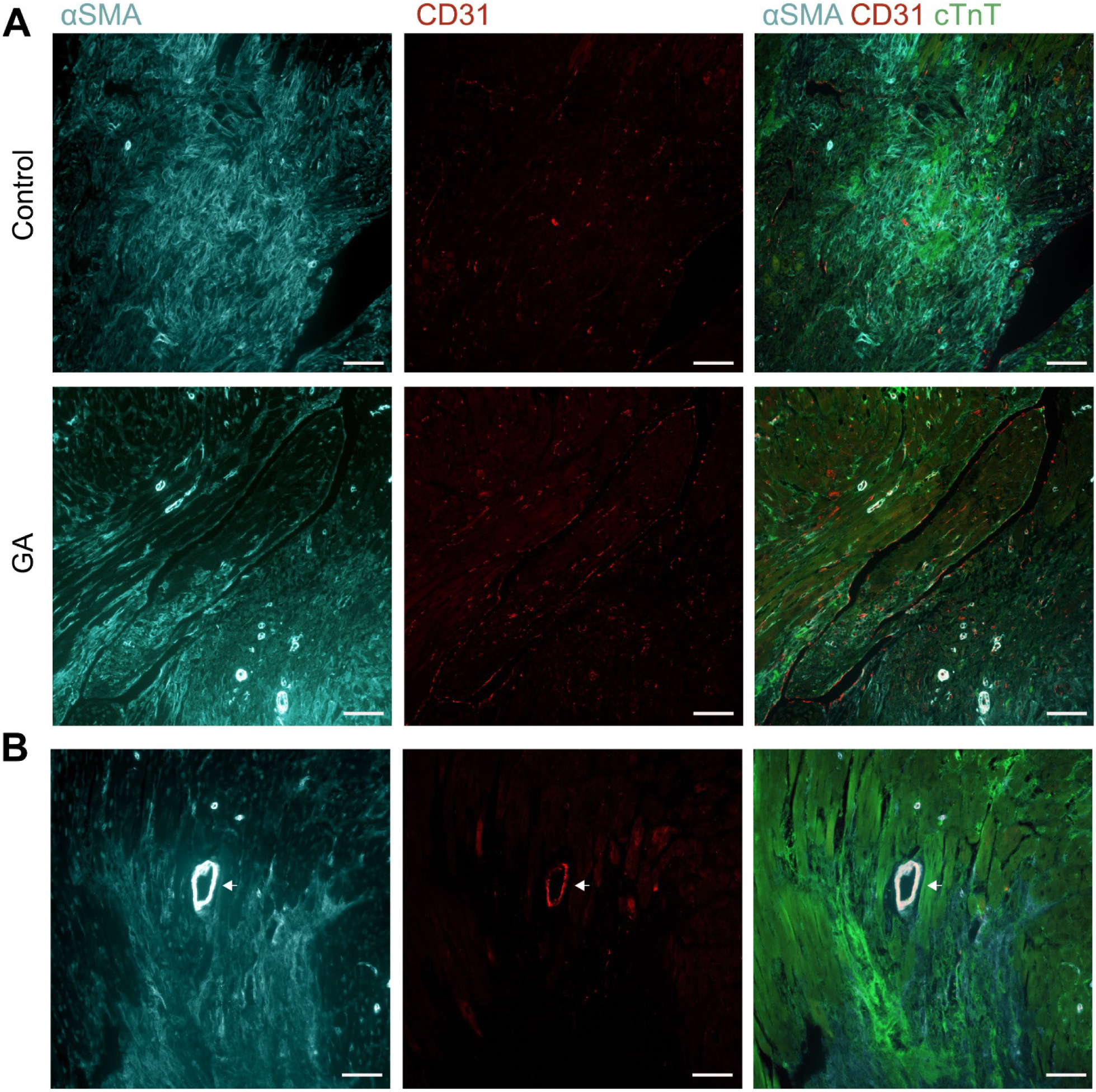
CD31+SMA+ colocalize in the walls of blood vessels. **A**, Representative images of cardiac sections, at 4 dpi of control (upper row) or GA (lower row). SMA+ areas (*left*) representing activated fibroblasts, adjacent to arterial blood vessels (circular structures), which are stained positive also for CD31+ (*right*). Webb-like structures staining solely for CD31+ (*middle*) represent capillaries. Scale bars: 400μm. **B**, A higher magnification of a SMA+CD31+ blood vessel, representing an artery. Scale bars: 200μm.

**Figure S6:**
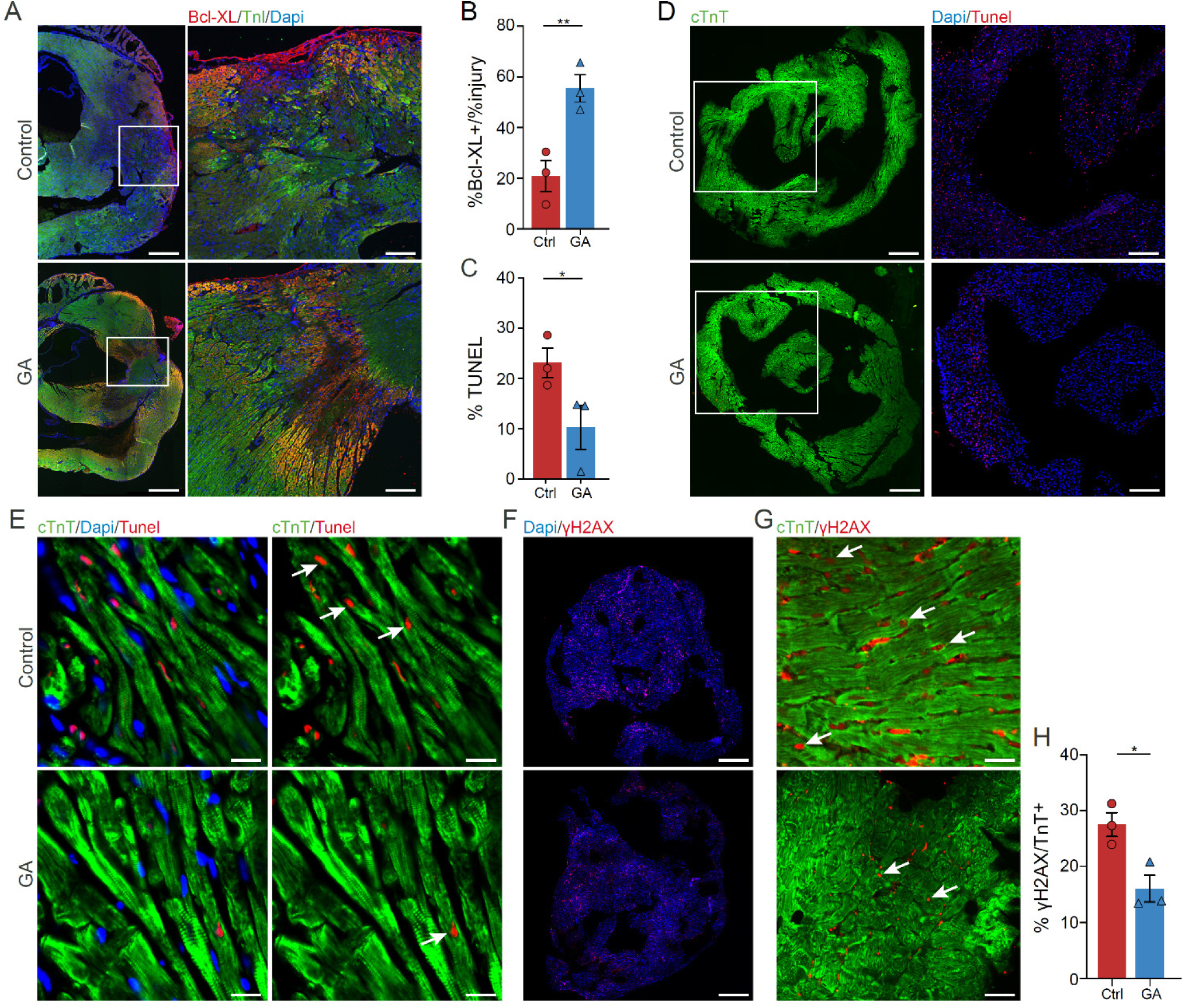
Glatiramer acetate protects cardiomyocytes from stress induced cell death *in vivo* and *ex vivo*. **A,** Representative images of cardiac sections of control and GA treated mice, at 1 dpi, stained for the anti-apoptotic protein, BcL-X_L_. Scale bars: left panels, 1000μm; right panels, 250μm. **B,** Quantification of %BcL-X_L_ area in the LV, normalized to the % of injured area (n=3 for each group). **C,** Quantification of TUNEL positive cells in control and GA treated EVOC (n=3 for each group. **D,** Representative images showing TUNEL positive cells in control and GA treated EVOC that were exposed to H_2_O_2_. Scale bars: 500μm. **E,** Higher magnifications of D, showing CMs positive for TUNEL. Scale bars: 20μm. **F,** Representative images of control and GA treated EVOC that were exposed to H_2_O_2_ and stained for the DNA double-strand breaks marker γH2AX. Scale bars: 500μm. **G,** Higher magnifications of F, showing CMs positive for γH2AX. Scale bars: 20μm. **H,** Quantification of % γH2AX positive cells (n=3 for each group, an average of 550 cells were counted for each group).

**Fig S7:**
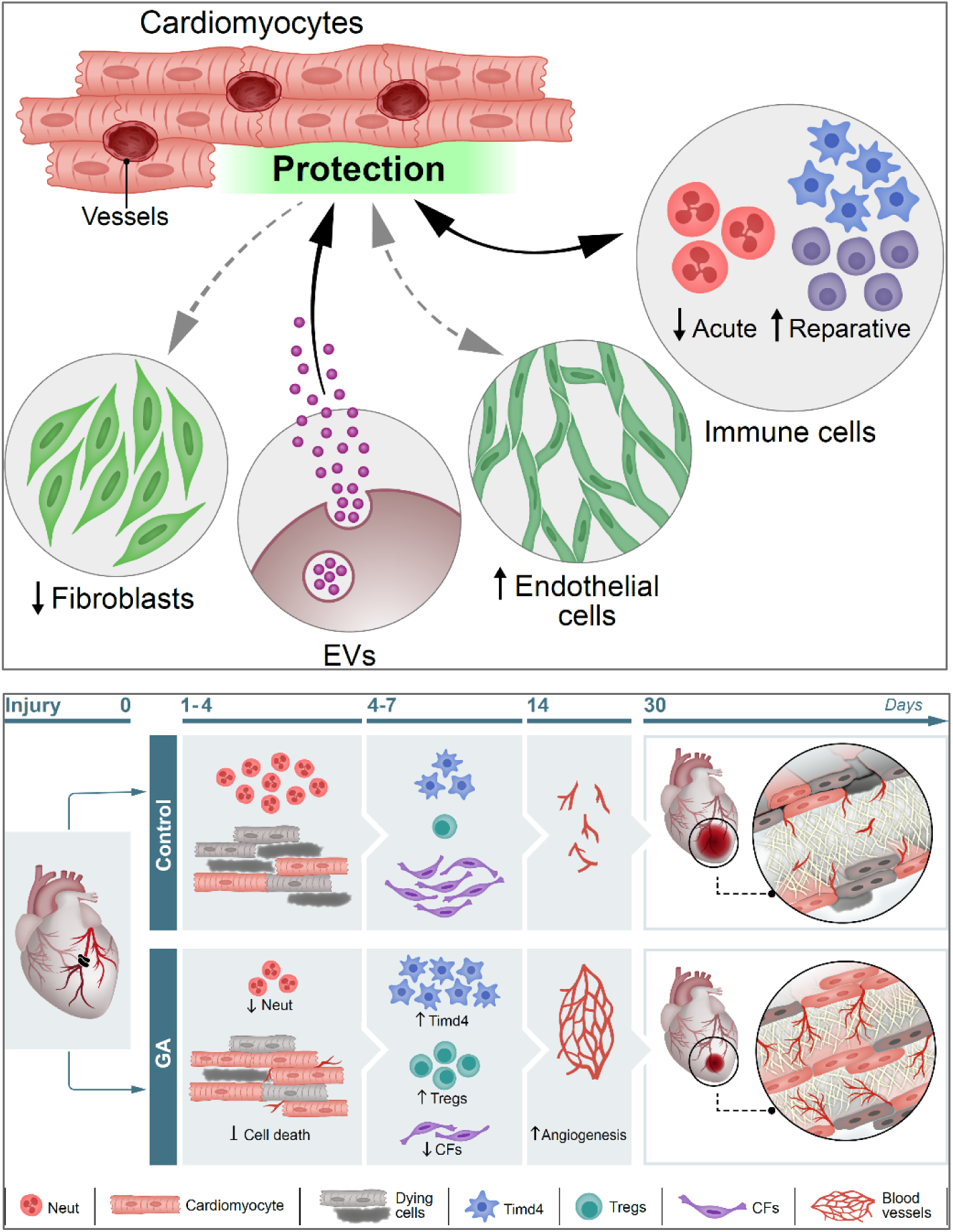
Models summarizing the pleiotropic beneficial effect of GA on the injured heart. Illustrations summarizing the effect of GA on multiple cardiac cell types and the crosstalk between them (upper). Black and grey arrows represent direct and indirect effects, respectively. The effect of GA on distinct cell populations is demonstrated by the time course of the cells’ participation in the repair process (lower).

**Figure S8:**
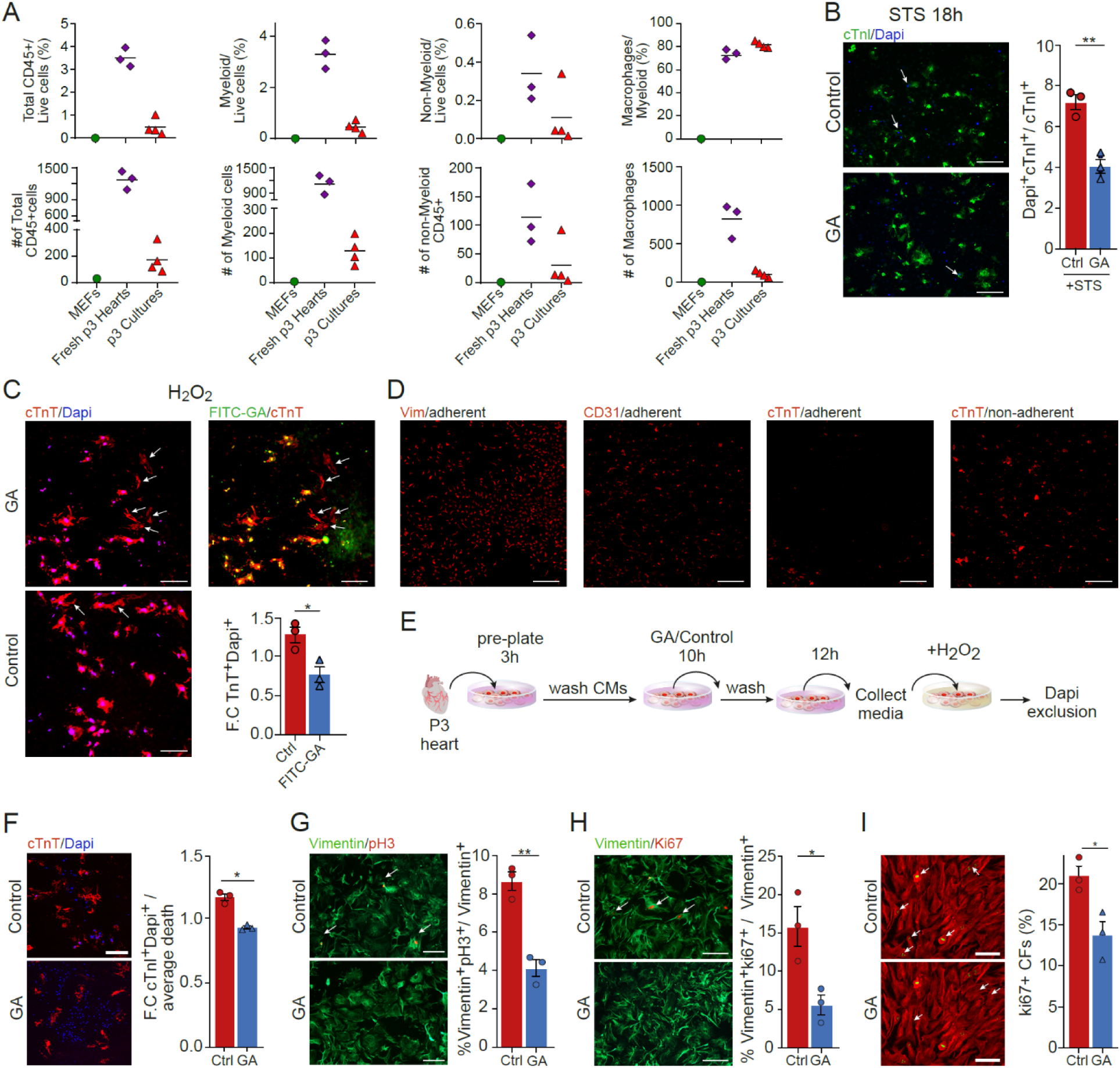
The effects of GA on mouse and human cultured cardiac cells. **A,** FACS analysis of P3 cultures used for the *in vitro* experiments reveals negligible amounts of immune cells in the cultures. **B,** P3 cultures were treated either with STS alone or with STS + GA. Apoptotic CM death was measured by DAPI exclusion assay 18h after the addition of STS. Arrows point to dying cells (n_PBS_ = 4, 700 cells; n_GA_ = 5, 1321 cells). Scale bars: 300 μm. **C,** P3 cultures were treated with FITC-GA for 48h, followed by treatment with 1 µM H_2_O_2_ for 2h. DAPI exclusion assay was performed to determine the levels of cell death, followed by staining with anti-cTnT (red). Arrowheads point at cTnT+DAPI-cells, representing live CMs in GA-treated (top) and control cultures (bottom left). Bottom right panel shows the quantification of the fold change in cTnT+DAPI+ dead CMs between the two groups (n=3 for each group). In total, 1248 cells were counted. Scale bars: 200 μm. **D,** Representative images of adherent cells at 3h cultures, showing that most of the cultures contain CFs (vimentin) and ECs (CD31), whereas CMs (cTnT) are mainly non-adherent at this time point. Scale bars: 750 μm. **E,** A scheme showing the experimental design of the conditioned media assay. **F,** DAPI exclusion assay following challenge with H_2_O_2_ and treatment with conditioned medium. Quantification of the fold-change in cTnI+DAPI+ (right) (n=3, 1487 cells). Scale bars: 300 μm. **G,H,** P3 cultures were treated with either GA or control for 48h and co-stained with anti-vimentin and anti-pH3 (G) or anti-vimentin and Ki67 (H). Quantifications show less proliferation of CFs compared to control (n=3 for each group). In total, 2482 and 3616 cells were counted for GA and PBS, respectively. Scale bars: 200 μm. **I,** Representative fields (left) and quantification (right) show less proliferation of CFs in GA-treated cultures derived from human biopsies. Scale bars: 150 μm. (n = 3, more than >3000 cells for each group).

**Figure S9:**
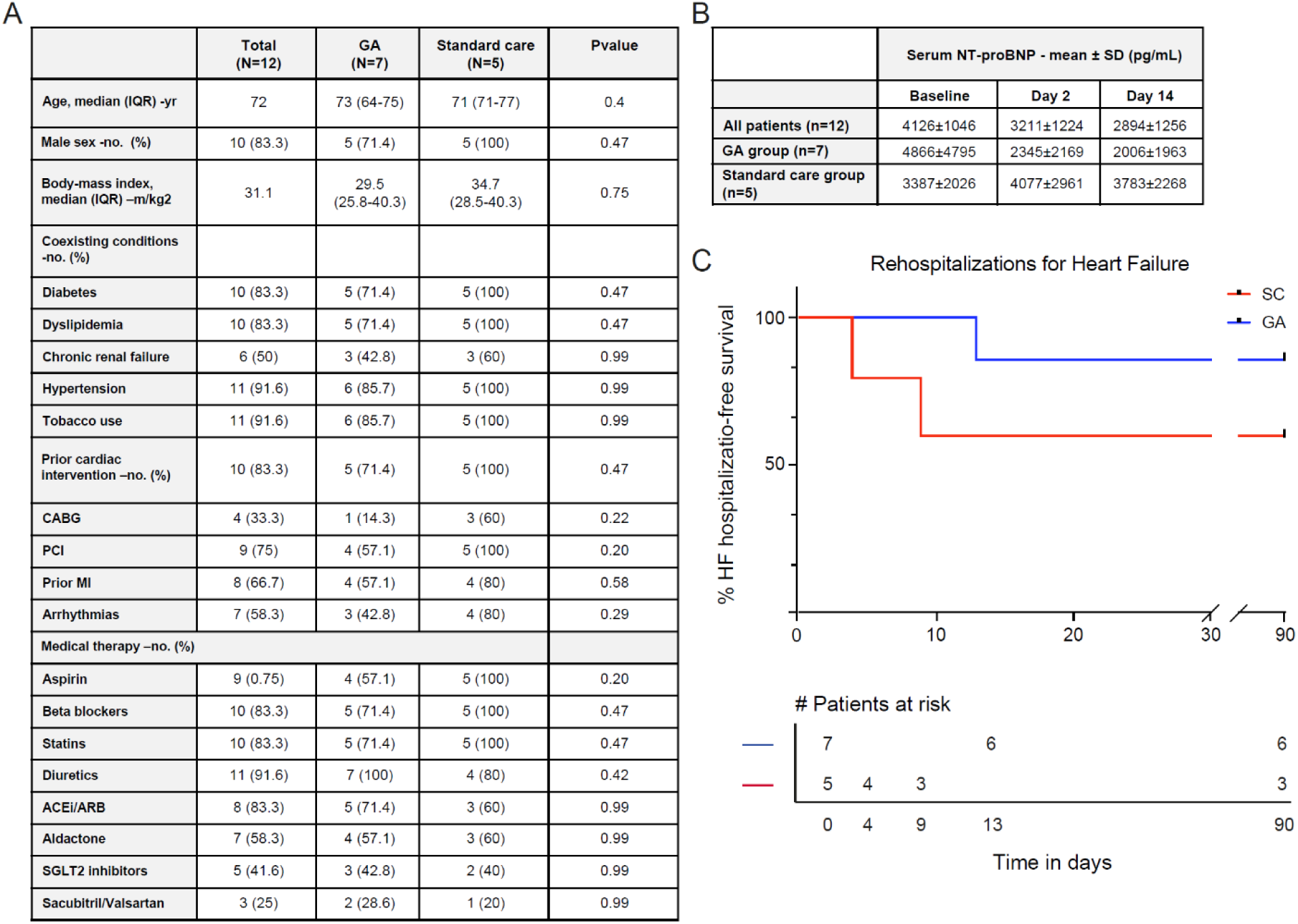
Additional data of the patients that participated in the clinical trial. **A**, An extended list of baseline patient characteristics, including demographics, cardiovascular co-morbidities and medical therapy. Continuous variables are presented as median and interquartile range (IQR), discrete variables as proportions. **B,** Raw values of serum NT-proBNP in the entire cohort according to treatment group. NT-proBNP were markedly decreased already at 2 days after initiation of GA therapy, while the opposite trend was observed in the control group. 14 days after hospitalization, NT-proBNP levels were close to baseline levels in the control group, but still markedly lower in the GA-treated group. **C,** Kaplan-Meier curve showing HF hospitalization-free in the 3 months following enrollment. Although the number of re-hospitalizations was small (n=3), the relative reduction in risk of rehospitalization in the GA-treated group is 64%.

**Figure S10:**
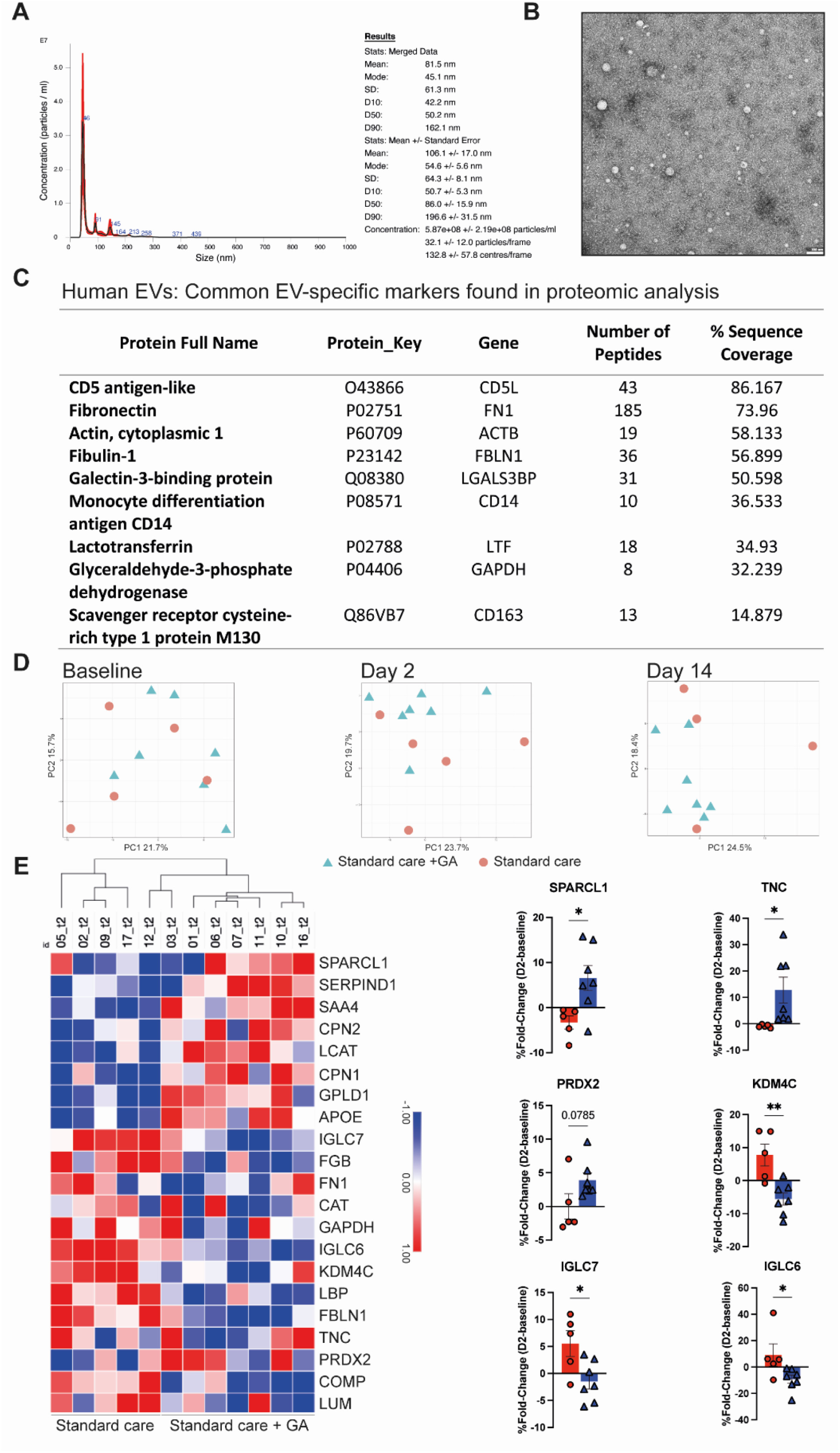
Proteomic analysis of EV-enriched serum from patients with ADHF. Proteomic analysis was performed using mass spectrometry on sera from patients that participated in the clinical trial. The sera were enriched for EVs using ultracentrifugation. **A**, Nanoparticle tracking analysis, demonstrating that the majority of particles detected are below 200 nm, averaging 106 nm, characteristic of EVs. **B**, Transmission electron microscopy. Samples were stained with 2% uranyl acetate, revealing multiple vesicles. Scale: 100 nm. **C**, Proteomic analysis showing characteristic protein markers for EVs, suggestive of an EV-enriched serum. **D**, Principal component analysis at three timepoints: baseline, day 2 and 14. At baseline all samples are scattered randomly. Starting at day 2, a separate cluster of GA-treated patients is apparent, suggesting a common set of enriched proteins. **E**, A heat map of day 2, generated from DE proteins in ANOVA, using mean intensities transformed on log2. Dendrogram showing clustering according to treatment, and a distinct pattern of proteome expression is evident. **F**, Individual proteins that were found to be differentially expressed at day 2, shown as fold-change from baseline levels.

## Notes

### Competing Interest Statement

The authors have declared no competing interest.

### Clinical Trial

NCT06003972

### Funding Statement

179BThe study was supported by the following funding sources:
180BEuropean Research Council, ERC AdG grant no. 788194, CardHeal (E.T)
181BERC-PoC no. 899224, ReDHeaD (E.T)
182BEU Horizon 2020 Research and Innovation Programme REANIMA

### Author Declarations

Institutional Ethics committee and national ethics committee of Israeli Ministry of Health gave their ethical approval for this work.

